# BCGNet: An AI Model Trained on 600K Hours of Sleep Data for a Novel Under-Pillow Contactless Monitoring Device

**DOI:** 10.1101/2025.08.23.25334218

**Authors:** Shigeng Chen, Xuesong Chen, Weijun Huang, Fei Lei, Chuxuan Shan, Zengrui Jin, Yunhan Shi, Yichen Wang, Rui Zhao, Xing Xu, Dongsheng Lv, Yanru Li, Shirui Pan, Ambrose Chiang, M. Brandon Westover, Shenda Hong, Chao Zhang, Shankai Yin, Chun-feng Liu, Hongliang Yi, Xiangdong Tang, Yue Leng

## Abstract

Sleep profoundly impacts health, yet current gold-standard Polysomnogram (PSG) is constrained by cost, discomfort, and limited scalability for longitudinal monitoring. Ballistocardiogram (BCG) offers a non-invasive and user-friendly alternative but often lacks the precision needed for reliable real-world applications. To address this gap, we propose BCGNet, a two-stage transfer learning model that is first pre-trained on 580,865 hours of PSG and then fine-tuned and validated on 15,081 hours of BCG (total 595,946 hours of recordings). Across multiple validation cohorts, BCGNet achieves strong performance in 4-class sleep staging (F1: 0.710 − 0.817), Apnea-Hypopnea Index (AHI3%) estimation (Pearson’s r > 0.95), and robust quantification of sleep continuity and architecture (ICC and Pearson’s r generally > 0.8). Notably, BCGNet maintains strong performance even on short daytime naps and demonstrates excellent generalizability across diverse external datasets. Deployed as a portable, contactless sleep tracking mat, BCGNet represents a major step towards scalable, user-friendly solutions for longitudinal home sleep monitoring, with important implications for population screening and personalized sleep medicine.

## 1 Introduction

Sleep profoundly impacts daily health, with poor sleep consistently linked to increased risks of cardiovascular, metabolic, and neuropsychiatric disorders [1–4]. As such, sleep monitoring is essential not only for diagnosing sleep disorders but also for enabling early detection of broader health conditions. Currently, Polysomnogram (PSG) remains the gold standard for evaluating sleep architecture and diagnosing sleep-related disorders [5–7]. However, PSG requires overnight monitoring in a sleep laboratory with multiple sensors attached to the body, which can cause discomfort and introduce first-night effects that may alter sleep patterns [8, 9]. Moreover, standard PSG protocols typically capture only a single night of data, limiting the ability to assess night-to-night variability in habitual sleep patterns [10]. These factors—limited comfort, high cost, and lack of longitudinal capability—make PSG poorly suited for continuous, long-term monitoring in home settings [11, 12].

Ballistocardiogram (BCG) has emerged as a promising non-invasive approach for sleep monitoring, capturing the subtle ballistic forces generated by cardiac ejection, respiratory excursions, and gross body movements during sleep [13, 14]. While BCG offers significant advantages in comfort and feasibility for at-home use [15–19], its reliance on mechanical vibrations makes it highly susceptible to motion artifacts and ambient noise, compromising its reliability in real-world environments [20–23]. This creates a critical unmet need: a technology that combines the diagnostic precision of PSG with the comfort and scalability of home-based sensors.

To address this challenge, we propose a novel transfer learning method that bridges the gap between these two modalities. We hypothesized that a model pre-trained on rich, high-fidelity data from laboratory-grade PSG could be fine-tuned to interpret the sparser, noisier signals of BCG—effectively enabling the model to “learn” complex physiological patterns within the simpler BCG input by transferring knowledge from PSG. To achieve this, we developed BCGNet (Figure 1(c)), a specialized two-stage model based on 580,865 hours of PSG recordings and 15,081 hours of BCG recordings, to robustly translate diagnostic features from the PSG domain into the BCG domain.

**Fig. 1.**
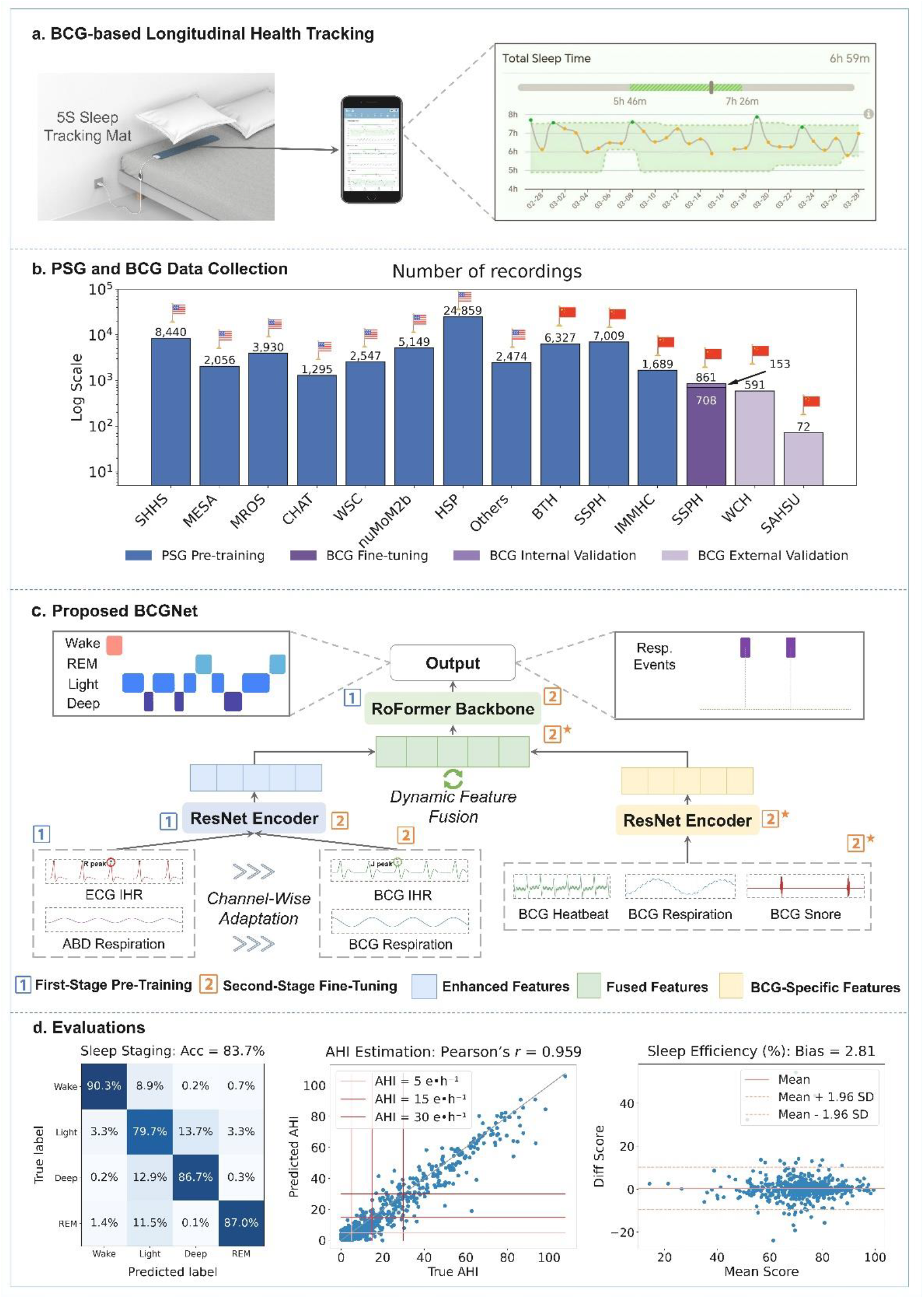
Overview of the proposed BCGNet and its validation. **(a)** Our method of BCG-based longitudinal sleep tracking using the 5S Sleep Tracking Mat. The in-mat sensor collects sleep data, and results are visualized on a mobile application for daily monitoring. **(b)** The scale and composition of datasets used in this study. The PSG cohorts were from the U.S. and China (noted by the flags). The “Others” included more datasets from NSRR (see Section 4.1) to diversify and enrich pre-training data. The BCG SSPH was divided such that 708 recordings were utilized for fine-tuning, and 153 recordings were reserved for internal validation. The WCH and SAHSU cohorts served as external cohorts. All dataset splits were performed strictly at the patient-level to avoid data leakage (best viewed in color). **(c)** The architecture of the proposed BCGNet, which first learns robust representations from ECG and ABD signals, then fine-tunes on BCG signals for final predictions. **(d)** Representative evaluation results on the WCH cohort. From left to right: a confusion matrix for 4-class sleep staging, a scatter plot for AHI3% estimation, and a Bland-Altman plot for sleep efficiency evaluation.

To the best of our knowledge, we are the first to apply transfer learning to bridge the gold-standard PSG—specifically its electrocardiogram (ECG) and abdominal (ABD) channels—to the contactless BCG. We validated our BCGNet in both internal and external cohorts, confirming its strong performance in key sleep assessment domains, including 3-stage (Wake/NREM/REM), 4-stage (Wake/N1+N2/N3/REM, reported in the main text), and 5-stage (Wake/N1/N2/N3/REM) classification; estimation of the Apnea– Hypopnea Index (AHI3%, as defined in the *Recommended rule 1A* according to the American Academy of Sleep Medicine (AASM) [24] criteria, with hypopneas scored if they were associated with either a ≥ 3% oxygen desaturation or an arousal); and the quantification of detailed sleep architecture during both overnight sleep and short daytime naps. Importantly, the algorithm exhibits strong generalizability and reliability across external cohorts, marking a significant step towards clinically robust, contactless sleep assessment.

While under-mattress devices such as Withings [25] and Sleeptracker-AI [26] have shown feasibility for home OSA screening, most existing systems still face challenges in balancing signal fidelity and long-term reliability with maintaining user-imperceptible operation in real-world settings. In this work, we use the Five Seasons (5S) Sleep Tracking Mat as the BCG acquisition device, as shown in Figure 1(a). Placed under the pillow rather than under the mattress, the 5S mat provides a user-friendly, fully contact-free interface suitable for routine use in both clinical and home environments (details in Section 4.2). Using this platform, we collected 15,081 hours of longitudinal BCG recordings, which form the BCG dataset used to develop and validate BCGNet and to demonstrate its potential for scalable OSA screening and long-term, contactless sleep monitoring in real-world conditions.

## 2 Results

### 2.1 Overview of the BCGNet

As illustrated in Figure 1(c), our BCGNet harnesses rich pretrained representations through a two-stage transfer learning process from PSG signals. Specifically, during first-stage pre-training, Instantaneous Heart Rate (IHR) and respiratory signals, derived respectively from ECG and ABD channels of PSG recordings, are encoded via a ResNet [27] encoder to generate enhanced features. These features are subsequently processed through a RoFormer [28] backbone to capture temporal variations. In the second-stage fine-tuning, BCG-derived IHR and respiratory signals are fed into the model layers pretrained on PSG to enable *Channel-Wise Adaptation*. Concurrently, discriminative BCG-specific features, captured via bandpass filtering (marked with asterisks), are adaptively combined with the enhanced features using a *Dynamic Feature Fusion* technique. The fused representation is passed through the RoFormer backbone to generate the final output. Implementation and training details of BCGNet are described in Section 4.4.

During inference, the model relies solely on BCG, including IHR, respiratory signals, and BCG-specific features, for predicting sleep stages and respiratory events. This design allows the model to operate independently of PSG data at deployment, enabling practical, real-world applications where only BCG signals are available.

### 2.2 Study population

Table 1 summarizes the demographic and anthropometric characteristics of the main cohorts used in this study. A key strength of our study is the unprecedented scale and diversity of the data collected. In total, we collected 595,946 hours of sleep recordings from 53,544 subjects to develop BCGNet. For the pre-training stage, we aggregated 580,865 hours of polysomnography (PSG) recordings drawn from over ten diverse cohorts. These included large-scale, publicly available U.S. datasets such as the

**Table 1:** Summary of datasets.

| Dataset | Subjects | Recordings | Hours | Female | Age | BMI |
| --- | --- | --- | --- | --- | --- | --- |
| **PSG<sup>†</sup>** |  |  |  |  |  |  |
| SHHS | 5,795 | 8,440 | 75,432 | 52.8% | 64.5 ± 22.3 | 28.2 ± 10.2 |
| MESA | 2,056 | 2,056 | 21,745 | 53.6% | 69.4 ± 18.2 | 28.7 ± 11.2 |
| MROS | 2,905 | 3,930 | 45,110 | 0.0% | 77.6 ± 11.2 | 27.1 ± 7.6 |
| CHAT | 1,006 | 1,295 | 12,884 | 26.4% | 7.0 ± 2.9 | 19.1 ± 9.8 |
| WSC | 1,105 | 2,547 | 20,334 | 46.1% | 59.8 ± 17.0 | 31.7 ± 14.4 |
| nuMoM2b | 3,114 | 5,149 | 59,795 | 100.0% | 27.0 ± 11.0 | 27.6 ± 12.8 |
| HSP | 18,701 | 24,859 | 187,761 | 43.0% | 53.1 ± 33.4 | 31.6 ± 15.3 |
| BTH | 6,327 | 6,327 | 58,639 | 22.7% | 34.5 ± 17.8 | 24.8 ± 5.4 |
| SSPH | 7,009 | 7,009 | 61,504 | 20.9% | 40.5 ± 25.8 | 26.3 ± 13.2 |
| IMMHC | 1,689 | 1,689 | 12,982 | 63.6% | 52.0 ± 30.4 | 24.1 ± 7.7 |
| Others | 2,313 | 2,474 | 24,679 | 58.9% | 38.8 ± 56.1 | 27.9 ± 18.6 |
| **BCG** |  |  |  |  |  |  |
| SSPH<sup>‡</sup> | 708 | 708 | 6,960 | 27.7% | 43.5 ± 24.9 | 26.1 ± 7.7 |
| SSPH<sup>§</sup> | 153 | 153 | 1,522 | 36.3% | 42.0 ± 26.4 | 24.4 ± 7.1 |
| WCH<sup>§</sup> | 591 | 591 | 5,872 | 25.8% | 42.8 ± 30.8 | 25.9 ± 12.8 |
| SAHSU<sup>§</sup> | 72 | 72 | 727 | 18.1% | 44.5 ± 32.5 | 26.6 ± 8.1 |

Subjects = number of participants; Hours = total recording duration in hours; Age and BMI are presented as Mean ± 2SD.
<sup>†</sup>: Pre-training set. <sup>‡</sup>: Fine-tuning set. <sup>§</sup>: Validation sets. Note that all PSG cohorts were used exclusively for pretraining, and the BCG SSPH cohort (N=861) was split at the patient-level into a fine-tuning set (N=708) and an internal validation set (N=153). The WCH and SAHSU served as external validation cohorts.
**Full names of datasets:** SHHS = Sleep Heart Health Study; MESA = Multi-Ethnic Study of Atherosclerosis; MROS = Osteoporotic Fractures in Men Study; CHAT = Childhood Adenotonsillectomy Trial; WSC = Wisconsin Sleep Cohort; nuMoM2b = Nulliparous Pregnancy Outcomes Study: Monitoring Mothers-to-Be; HSP = Human Sleep Project; BTH = Beijing Tongren Hospital; SSPH = Shanghai Sixth People’s Hospital; IMMHC = Inner Mongolia Mental Health Center; WCH = West China Hospital; SAHSU = The Second Affiliated Hospital of Soochow University. “Others” refers additional datasets from NSRR repository, including the Cleveland Family Study (CFS), Study of Osteoporotic Fractures (SOF), Cleveland Children’s Sleep and Health Study (CCSHS), Pediatric Adenotonsillectomy Trial of Snoring (PATS), and Home Positive Airway Pressure (HomePAP).

Human Sleep Project (HSP) and the National Sleep Research Resource (NSRR), together with hospital-based cohorts in China, including Beijing Tongren Hospital (BTH), Shanghai Sixth People’s Hospital (SSPH), and Inner Mongolia Mental Health Center (IMMHC) (see Supplementary Table 1 for sleep characteristics of the Chinese cohorts). The inclusion of cohorts with psychiatric-related conditions such as depression or insomnia further enriched the dataset, ensuring exposure to a wide spectrum of sleep phenotypes and clinical conditions, thereby enhancing overall diversity.

For the fine-tuning and validation stages, we employed 15,081 hours of BCG recordings from 1,524 subjects across three independent hospital cohorts in China. These data were acquired with time-synchronized PSG, and the ground-truth labels were derived from expert annotations in accordance with AASM guidelines. Specifically, a second SSPH cohort (N=861; 8,482 hours) was divided into a fine-tuning set (N=708; 6,960 hours) and an internal validation set (N=153; 1,522 hours). External validation was further performed using data from West China Hospital (WCH; N=591; 5,872 hours) and the Second Affiliated Hospital of Soochow University (SAHSU; N=72; 727 hours). To ensure rigorous assessment of model generalizability, strict patient-level separation was enforced, with no overlap among the pre-training, fine-tuning, and validation cohorts.

### 2.3 BCGNet precisely classifies various sleep stages

Our BCGNet model demonstrates robust performance in 4-class sleep staging across multiple independent cohorts, as summarized in Table 2 and visualized by the confusion matrices in Figure 2. The internal validation on the SSPH cohort yields the highest agreement, achieving a Cohen’s *κ* of 0.766 (95% CI: 0.750, 0.781) and an F1 of 0.817 (95% CI: 0.804, 0.831). Regarding external validation, the model maintains strong performance across diverse populations. Specifically, BCGNet achieves a Cohen’s *κ* of 0.736 (95% CI: 0.728, 0.743) and F1 of 0.710 (95% CI: 0.700, 0.718) in the WCH, and a Cohen’s *κ* of 0.697 (95% CI: 0.666, 0.725) and F1 of 0.781 (95% CI: 0.759, 0.800) in the SAHSU. The 3-class and 5-class sleep staging results can be found in Supplementary Figure 1-2, and the sensitivity and specificity are also reported in Supplementary Table 2-4. This multi-center validation underscores the generalizability and robustness of BCGNet for reliable sleep staging across these diverse cohorts.

**Table 2:**
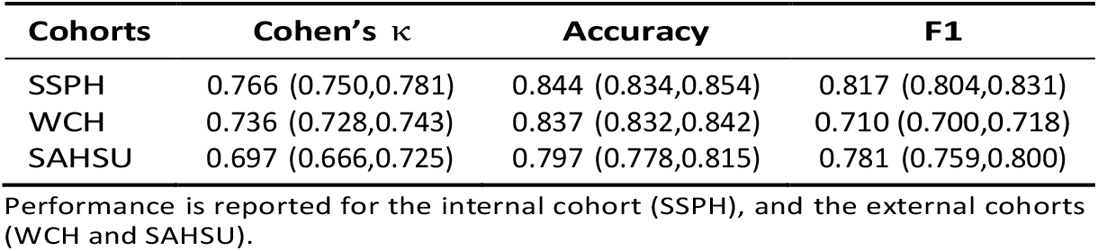
Sleep staging performance across multi-center cohorts.

**Fig. 2.**
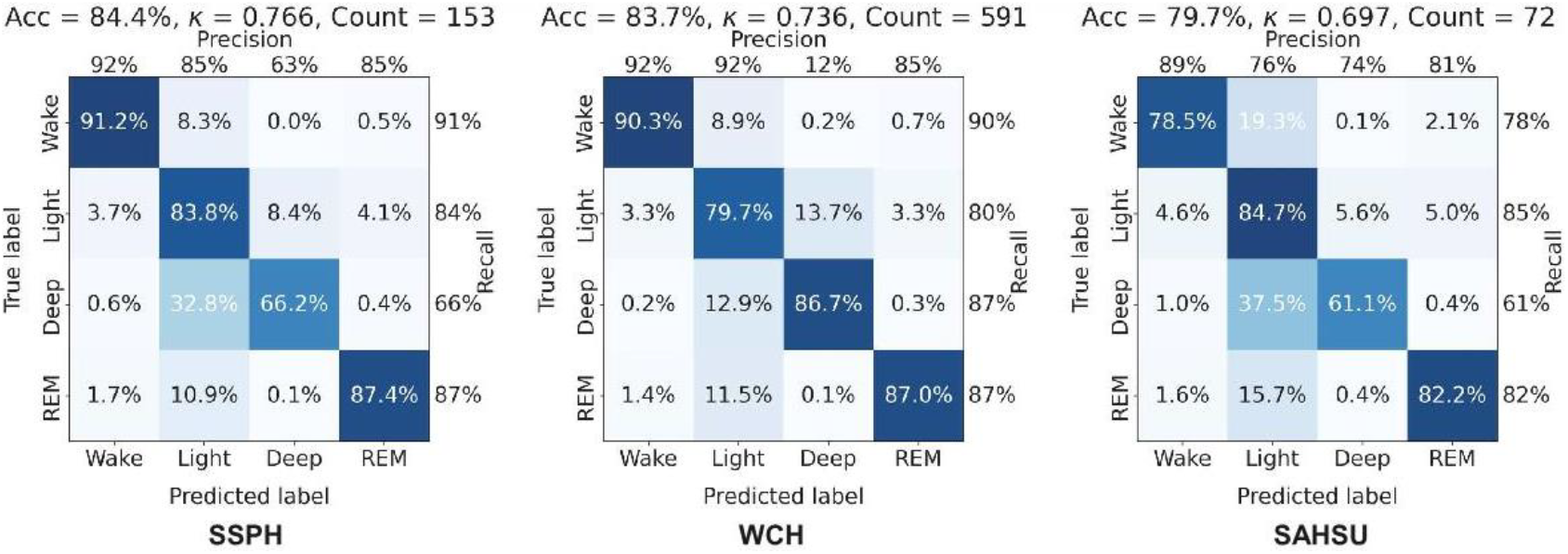
Performance of BCGNet for 4-class sleep staging across multi-center cohorts. Performance is reported for the internal validation cohort (SSPH) and the external validation cohorts (WCH and SAHSU). Overall accuracy (Acc), Cohen’s kappa (*κ*), and per-class precision and recall are provided above each matrix.

### 2.4 BCGNet robustly estimates AHI3% across multi-center cohorts

For continuous AHI3% estimation, the scatter plots in Figure 3 reveal a remarkably strong linear correlation between predicted and true AHI3% values across all cohorts. Pearson’s *r* is 0.963 for the SSPH cohort and remains high at 0.959 and 0.952 for the WCH and SAHSU external cohorts, respectively. The Bland-Altman plots in Figure 4 further reveal a strong level of agreement using the proposed BCGNet, with minimal mean biases (SSPH: -1.63 events/h; WCH: -2.35 events/h; SAHSU: 0.54 events/h) and tight 95% limits of agreement (LoA) (SSPH: [-11.74, 8.49]; WCH: [-15.11, 10.00]; SAHSU: [-13.01, 14.09]). This overall low bias and strong concordance across all cohorts, both internal and external, highlight the model’s robust calibration for AHI3% prediction in diverse clinical settings.

**Fig. 3.**
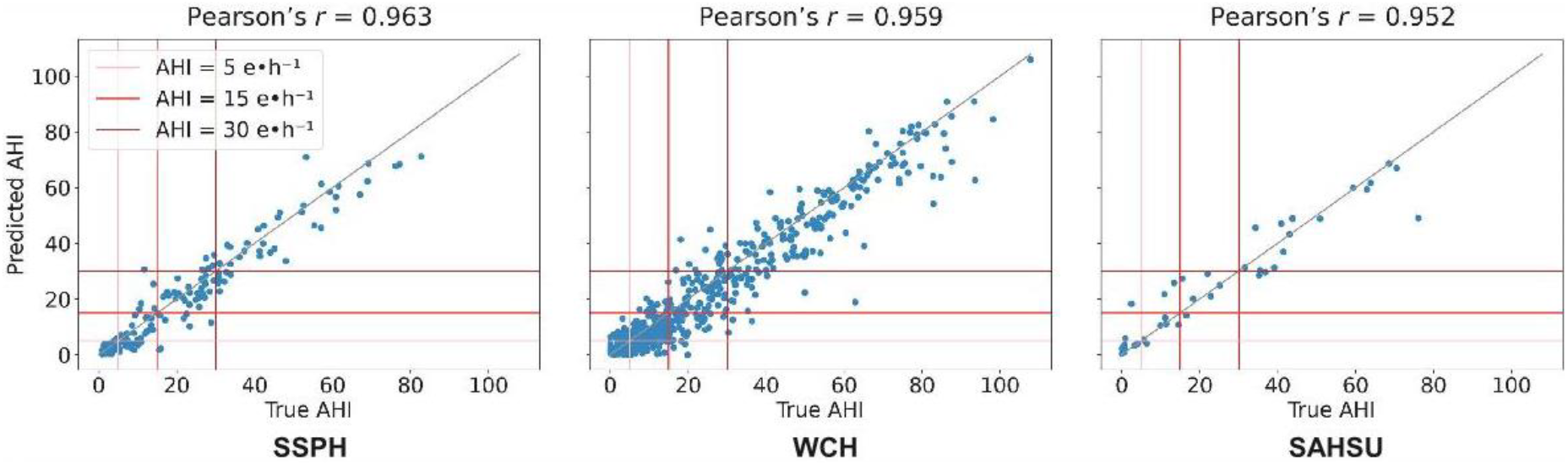
Scatter plots for AHI3% estimation. Performance is reported for the internal validation cohort (SSPH) and the external validation cohorts (WCH and SAHSU). These plots show predicted AHI3% versus true AHI3% values.

**Fig. 4.**
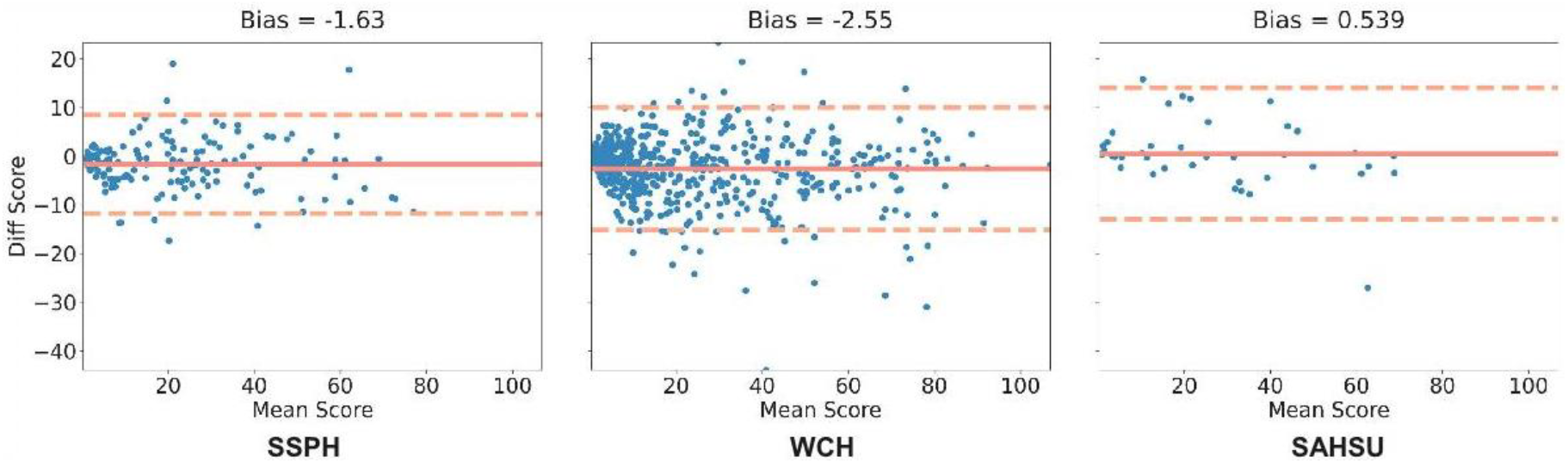
Bland-Altman plots for AHI3% estimation. Performance is reported for the internal validation cohort (SSPH) and the external validation cohorts (WCH and SAHSU). These plots illustrate the agreement between the predicted and true AHI3% values, depicting the mean bias and the 95% limits of agreement.

The BCGNet also achieves high accuracy in sleep apnea severity classification (Healthy, Mild, Moderate, Severe), as shown in Supplementary Figure 3. In the SSPH cohort, the highest values along the confusion-matrix diagonal, such as 94.3% for Healthy and 82.5% for Severe, demonstrate BCGNet’s strong classification performance across all categories. This robust performance is maintained in the external cohorts, which show high recall of 84.7% and 88.9% for Severe cases in the WCH and SAHSU, respectively. These results highlight BCGNet’s capability in identifying clinically significant sleep apnea across diverse populations. Furthermore, the bar charts in Supplementary Figure 4 demonstrate the model’s high diagnostic utility across standard AHI thresholds (5, 15, and 30 events/h). BCGNet consistently achieves high sensitivity and specificity for detecting mild, moderate, and severe sleep apnea levels across all validation cohorts. This robust and consistent performance at clinically meaningful cutoffs reinforces the model’s potential for accurate and reliable automated sleep apnea assessment in real-world applications.

### 2.5 BCGNet accurately measures overnight sleep continuity and architecture

Beyond sleep staging and AHI3% estimation, BCGNet also demonstrates strong performance in quantifying key sleep continuity parameters—Total Sleep Time (TST), Sleep Onset Latency (SOL), Wake After Sleep Onset (WASO), and Sleep Efficiency (SE)—as well as the architecture parameter REM Latency (REML) from full-night recordings.

As shown in Figure 5, the model consistently achieves high Intraclass Correlation Coefficient (ICC) and Pearson’s correlation coefficients (*r*) across all evaluated sleep parameters and cohorts. In the internal SSPH cohort, both ICC and Pearson’s *r* values for TST, SOL, WASO, SE, and REML generally exceed 0.8, with TST and SE showing particularly strong agreement (both ICC and *r >* 0.97). In the external cohorts, agreement remains high, with TST and SE correlations also exceeding 0.9. While a slight decrease in correlation is observed in the individual external WCH and SAHSU cohorts, the performance remains robust, with most ICC and *r* values exceeding 0.8, indicating strong reliability and linear association between predicted and ground-truth values.

**Fig. 5.**
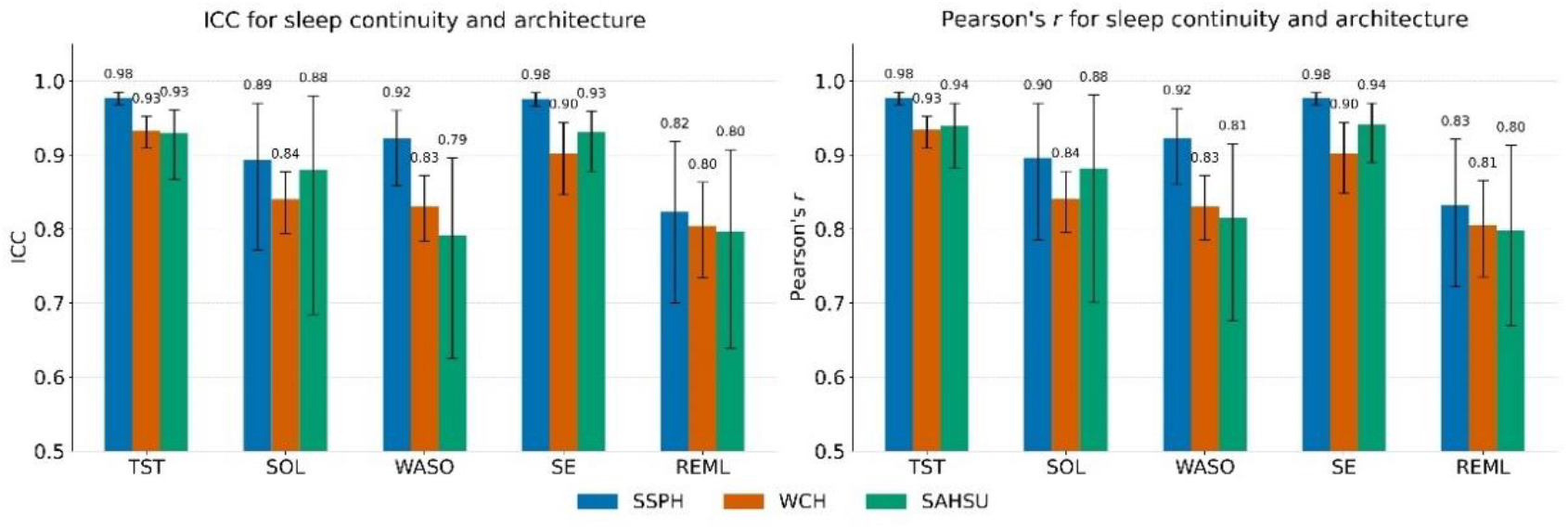
Performance of BCGNet in estimating overnight sleep continuity and architecture parameters. This figure presents the ICC and Pearson’s correlation coefficient (*r*) for five key sleep parameters: TST, SOL, WASO, SE, and REML. Results are grouped by parameter and displayed for three validation cohorts (SSPH, WCH, and SAHSU). Error bars represent the 95% confidence intervals.

The Bland-Altman plots in Supplementary Figure 5 provide a detailed visualization of the agreement between BCGNet predictions and expert annotations for each sleep parameter across the SSPH, WCH, and SAHSU cohorts. Across all parameters and cohorts, the plots consistently reveal minimal mean biases, as indicated by the solid red line close to zero, suggesting that model predictions closely align with ground-truth values. The 95% LoA, as indicated by dashed red lines, demonstrates that the majority of the data points fall within acceptable bounds, reflecting a high degree of precision and consistency. As expected, parameters with greater inherent variability, such as SOL and WASO, exhibited slightly wider LoA. In contrast, parameters like TST and SE showed tight clustering around the mean difference line, underscoring strong agreement.

### 2.6 BCGNet supports precise monitoring of daytime naps

Beyond the comprehensive assessment of overnight sleep, we further evaluated the performance of BCGNet on daytime naps. For this evaluation, we utilized data from a separate WCH cohort specifically dedicated to multiple sleep latency test (MSLT) studies. For daytime-nap analysis, we applied the same four-stage sleep-staging model used for nocturnal recordings and derived key sleep summary measures from its output. Given that short naps often contain few or no deep stages (N3, REM), we focused on three continuity-related metrics: TST, SOL, and SE at the patient level.

As presented in Table 6(a) and further illustrated by the 2-class sleep staging confusion matrix in Figure 6(b), our model demonstrated reliable performance for short naps. For the selected metrics, BCGNet achieved an ICC of 0.717 (95% CI: 0.663, 0.765) and a Pearson’s *r* of 0.725 (95% CI: 0.673, 0.773) for TST. SOL showed an ICC of 0.638 (95% CI: 0.578, 0.693) and a Pearson’s *r* of 0.653 (95% CI: 0.590, 0.712). Similarly, SE yielded an ICC of 0.669 (95% CI: 0.620, 0.709) and a Pearson’s *r* of 0.672 (95% CI: 0.623, 0.714). These results indicate a good to substantial agreement and correlation, confirming the model’s capability to quantify these essential parameters even in short sleep episodes. Furthermore, the Sleep-Wake staging for short naps highlights the model’s robust ability to differentiate between wakefulness and sleep states. With an overall accuracy of 81.3% and a Cohen’s *κ* of 0.624, the model achieved high precision (83% for Wake, 80% for Sleep) and recall (76% for Wake, 86% for Sleep). This strong binary classification performance is crucial for accurately determining the presence and duration of daytime naps.

**Fig. 6.**
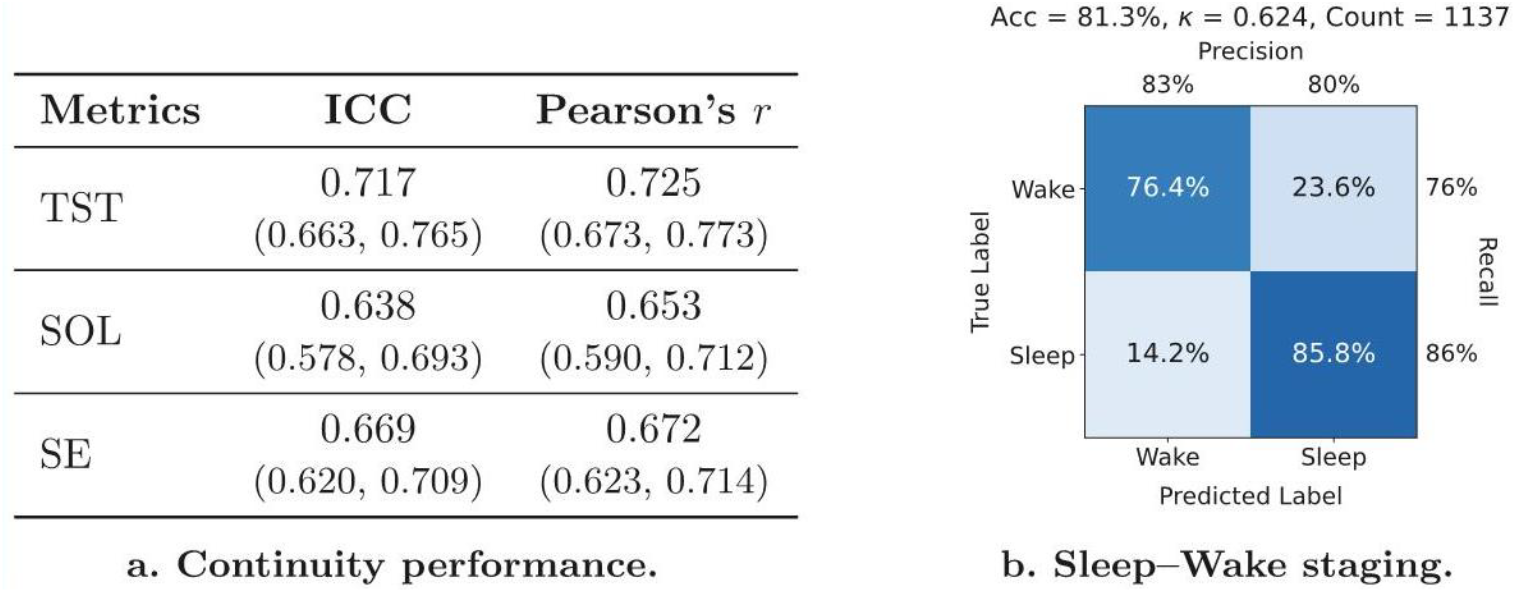
Evaluation of BCGNet performance on daytime naps. **(a)** For short naps, Total Sleep Time (TST), Sleep Onset Latency (SOL), and Sleep Efficiency (SE) are included as sleep continuity metrics. **(b)** Sleep–Wake staging evaluation is performed on the short-snap cohort.

In summary, BCGNet exhibits exceptional versatility and accuracy, extending its robust performance from comprehensive overnight sleep analysis to the nuanced assessment of daytime naps, making it a powerful tool for continuous, daily sleep monitoring across various settings.

### 2.7 Ablation study

Ablation experiments were conducted to elucidate the contribution of each key component within the proposed BCGNet to its overall performance. Specifically, we compared three distinct training schemes: (i) BCG-based, a model trained purely on BCG signals without PSG pre-training; (ii) PSG-based, representing a zero-shot condition where the PSG pre-trained model directly inferred on BCG data without fine-tuning; and (iii) Proposed BCGNet, our complete two-stage transfer learning model.

As presented in Table 3, the BCG-based model achieved a Cohen’s *κ* of 0.629 and an F1 of 0.717 for sleep staging. For AHI3% estimation, it yielded a Pearson’s *r* of 0.951 and an F1 of 0.713. The PSG-based model, while leveraging PSG-learned representations but not fine-tuning on BCG, showed reduced performance with Cohen’s *κ* of 0.742 and F1 of 0.795 for sleep staging, and Pearson’s *r* of 0.949 and F1 of 0.666 for AHI3% estimation. Notably, our proposed BCGNet generally demonstrated superior performance, achieving Cohen’s *κ* of 0.766 and F1 of 0.817 for sleep staging, and Pearson’s *r* (0.963) and F1 (0.711) for AHI3% estimation. Overall, these findings highlight that the two-stage training approach substantially improves the model performance by leveraging the transferred knowledge.

**Table 3:**
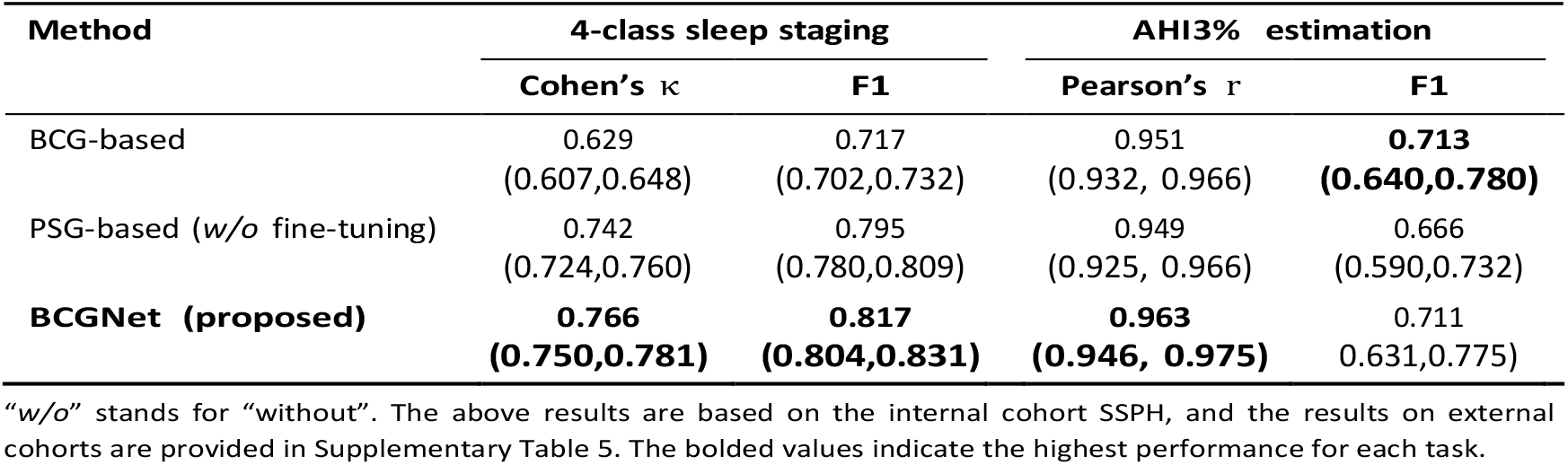
Results of ablation study on sleep staging and AHI estimation.

## 3 Discussion

In this study, we developed and validated BCGNet, a state-of-the-art transfer learning model enables comprehensive sleep assessment across multiple settings. Our findings demonstrate that BCGNet achieves high accuracy and robustness across multi-center cohorts for sleep staging, AHI estimation, assessment of key sleep continuity and architecture parameters, and analysis of short daytime naps. Compared to recent published studies (summarized in Table 4), our approach differs fundamentally in scale (595,946-hour recordings in total) and methodology, leading to superior performance and robustness. The proposed BCGNet not only outperforms previous BCG-based methods [33, 37] but also achieves performance comparable to or exceeding that of approaches based on PPG [29, 32], radio [36], ECG[30, 31, 34, 35], or pressure-sensitive sensors [25, 26]. A detailed comparison reveals several key advantages:

**Table 4:** Related work in the literature.

| Source (Year) | Sensor | Sleep staging | AHI estimation |
| --- | --- | --- | --- |
| Korkalainen et al. (2020) [29] | ◦ PPG (full signals) | 3-cl: Acc=0.80, $\kappa$=0.65<br>4-cl: Acc=0.69, $\kappa$=0.54<br>5-cl: Acc=0.64, $\kappa$=0.51 | - |
| Sun et al. (2020) [30] | ★ ECG, ABD (HR & Resp) | 5-cl: $\kappa$=0.53, F1=0.59 | - |
| Sridhar et al. (2020) [31] | ★ ECG (IHR) | 4-cl: Acc=0.72, $\kappa$=0.55 | - |
| Radha et al. (2021) [32] | ◦ PPG (HRV) | 4-cl: Acc=0.76, $\kappa$=0.65 | - |
| Li et al. (2024) [33] | ★ BCG (HR & Resp) | 5-cl: Acc=0.70, $\kappa$=0.59 | - |
| Liu et al. (2024) [34] | ◦ ECG (full signals) | - | cutoff at 5: Acc=0.81<br>cutoff at 15: Acc=0.91<br>cutoff at 30: Acc=0.93 |
| Zhang et al. (2025) [35] | ◦ ECG, SpO<sub>2</sub> (full signals) | - | cutoff at 5: Acc=0.90<br>cutoff at 15: Acc=0.88<br>cutoff at 30: Acc=0.92 |
| Edouard et al. (2021) [25] | ★ Withings<sup>α</sup> | - | cutoff at 5: Sens=0.89, Spec=0.75<br>cutoff at 15: Sens=0.88, Spec=0.89<br>cutoff at 30: Sens=0.86, Spec=0.91 |
| Ding et al. (2022) [26] | ★ Sleeptracker-AI<sup>β</sup> | 3-cl: Acc=0.87, $\kappa$=0.73<br>4-cl: Acc=0.79, $\kappa$=0.68 | cutoff at 5: Sens=0.86, Spec=0.88, Acc=0.87<br>15: Sens=0.82, Spec=0.93, Acc=0.92 |
| He et al. (2025) [36] | ★ Radio (wave) | 4-cl: Acc=0.84 | r=0.82<br>cutoff at 5: Sens=0.74, Spec=0.89<br>cutoff at 15: Sens=0.73, Spec=0.95 |
| Ours | ★ BCG (IHR & Resp) | 3-cl: Acc=0.91, $\kappa$=0.85, F1=0.90<br>4-cl: Acc=0.83, $\kappa$=0.73, F1=0.83<br>5-cl: Acc=0.74, $\kappa$=0.66, F1=0.67 | r=0.95, cutoff at 5: Sens=0.92, Spec=0.81, Acc=0.90<br>15: Sens=0.91, Spec=0.89, Acc=0.90<br>30: Sens=0.87, Spec=0.98, Acc=0.94 |

◦ evaluated through *internal* validation within the development dataset (e.g., via cross-validation or data splitting). ★ evaluated through *external* validation on an independent cohort distinct from the training cohort. <sup>α</sup> Withings is an under-mattress, pneumatic device that measures pressure changes in an air bladder. <sup>β</sup> Sleeptracker-AI is an under-mattress, piezo-electric monitor that measures forces exerted through the mattress. Results of “Ours” in this table are reported as the performance on the external validation cohorts (i.e., WCH and SAHSU).
“cls” stands for classification. For example, in sleep staging, “4-cl” refers to four-class staging; In AHI estimation, “4-cl” represents the four categories: healthy, mild, moderate, and severe. “full signals” refers to using the raw, continuous signals as the input to the model. “HR” and “Resp” refer to Heart Rate and Respiration, respectively. “HRV” and “IHR” are abbreviations for Heart Rate Variability and Instantaneous Heart Rate. For specific details on how these features are processed and extracted, please refer to the original papers.

Firstly, to our knowledge, the 5S Sleep Tracking Mat is the first under-pillow device for contactless sleep monitoring. In contrast to under-mattress systems [25, 26], our design prioritizes an enhanced, imperceptible user experience by placing the BCG sensor in the more challenging location beneath the head and torso. Although this placement results in more complex ballistocardiographic signals, it enables a superior contact-free monitoring solution with a user-centric design.

Secondly, the scale and diversity of our training data are unparalleled (see Figure 1(b) and Table 1). Previous studies on BCG-based sleep staging or apnea detection typically rely on smaller, single-center datasets, often numbering in the dozens or over one hundred subjects [37, 38]. Such models risk overfitting to local population characteristics and often fail to generalize beyond their training data [39, 40]. In contrast, our diverse and large training data exposed the BCGNet to a vast spectrum of sleep phenotypes, demographic variations (age, sex, BMI, ethnicity), and technical variabilities, providing it with a deep and generalizable understanding of sleep patterns that is foundational to its robustness.

Thirdly, we introduce a paradigm shift from direct training to transfer learning. Typically, transfer learning from PSG to BCG is challenging because BCG recordings exhibit a substantially lower signal-to-noise ratio (SNR) than PSG, which reduces the fidelity of directly transferred representations. However, our transfer learning model overcomes this limitation through a two-stage procedure. With *Channel-Wise Adaptation*, the model first learns robust, time-aware features from dual-channel PSG data (ECG and ABD). Subsequently, *Dynamic Feature Fusion* adaptively integrates these aligned high-level PSG-derived features with the raw continuous BCG-specific features from BCG, ensuring that the transferred information is both meaningful and usable despite the poorer SNR of BCG. As a result, our approach outperforms most existing models that are trained directly and exclusively on a single modality [41, 42].

More importantly, our validation is more comprehensive. We evaluate the model using metrics for both sleep staging and AHI estimation (see Table 4), as well as sleep architecture parameters (see Figure 5). In contrast, previous studies have typically focused on only a subset of these measures [31– 35, 43– 47]. Our thorough assessment demonstrates that BCGNet provides a broader, more complete, and more accurate solution for contactless sleep monitoring. Furthermore, validation on daytime naps broadens its applicability and potential for capturing a holistic picture of an individual’s 24-hour sleep-wake patterns, which are increasingly recognized as crucial for health and well-being [48, 49].

We acknowledge several limitations. First, the validation cohorts were composed of Chinese individuals referred for sleep studies. Future work aims to validate BCGNet on more ethnically and geographically diverse populations. Second, the assessment of short naps requires the user to be lying on the bed where the sleep monitoring mat is placed, which limits its application for tracking naps in other locations (e.g., on a sofa). Addressing this will require the development of portable or non-bed-based sensors capable of maintaining signal fidelity outside the traditional sleep environment. Additionally, although our model achieved high performance on the external cohorts, we observed a slight reduction in sleep staging when compared to its performance on the internal cohort. We speculate that this discrepancy may arise from the inherent variability in sleep stage scoring across different centers [50]. We also observed slightly lower performances for metrics such as SOL, WASO, and REML, particularly in the SAHSU cohort. Variation in performance across external cohorts is a known challenge in sleep monitoring studies, often attributed to differences in population characteristics and scoring protocols [51, 52]. While aggregate metrics like TST and SE are generally robust to minor scoring differences due to their reliance on macro-level sleep/wake patterns [51], parameters defined by precise transition points, such as SOL and WASO, are more sensitive to the inherent inter-scorer variability found between different sleep centers, particularly in the identification of Stage N1 and wake-to-sleep transitions [52, 53]. We therefore acknowledge this limitation as inherent to cross-cohort sleep assessment and plan to incorporate analysis based on annotations from multiple experts to better understand and mitigate the impact of such inconsistencies.

In conclusion, our BCGNet constitutes a notable methodological advance in contactless sleep monitoring. By effectively transferring knowledge from PSG to BCG signals, it achieves superior accuracy and coverage using entirely non-intrusive method. Our findings lay the groundwork for scalable, clinically validated tools that enable comfortable, continuous assessment of sleep health.

## 4 Methods

### 4.1 Datasets

We collected publicly available PSG datasets from multiple large-scale sleep studies to ensure diversity and robustness. Notably, all BCG recordings in SSPH, SAHSU, and WCH, were acquired with concurrent PSG recordings to provide gold-standard ground truths for model validation.

- **Human Sleep Project (HSP)** comprises 24,859 PSG recordings from 18,701 patients evaluated at the Massachusetts General Hospital Sleep Division, with signals acquired per AASM standards such as EEG, ECG, EOG, EMG, and airflow. The datasets are available at https://bdsp.io/content/hsp/2.0.
- **National Sleep Research Resource (NSRR)** provides a comprehensive repository of polysomnographic (PSG) data, demographic records, and clinical annotations from multicenter cohorts. We used the following main PSG subdatasets: Sleep Heart Health Study (SHHS), Multi-Ethnic Study of Atherosclerosis (MESA), Osteoporotic Fractures in Men Study (MROS), Childhood Adenotonsillectomy Trial (CHAT), Wisconsin Sleep Cohort (WSC), and Nulliparous Pregnancy Outcomes Study: Monitoring Mothers-to-Be (nuMoM2b). To enhance the model’s generalizability and robustness, we also included the following datasets: Cleveland Family Study (CFS), Study of Osteoporotic Fractures (SOF), Cleveland Children’s Sleep and Health Study (CCSHS), Pediatric Adenotillectomy Trial of Snoring (PATS), and Home Positive Airway Pressure (HomePAP). These datasets are obtained via https://sleepdata.org.
- **Shanghai Sixth People’s Hospital (SSPH affiliated to Shanghai Jiao Tong University School of Medicine)** contributed two distinct, non-overlapping datasets for this study. The first was a PSG-only dataset comprising 7,009 overnight recordings, used exclusively for the first-stage pre-training. The second was a dataset of 861 participants with simultaneously recorded PSG and BCG data; the BCG recordings from this latter cohort were used for the fine-tuning and internal validation stages. Both datasets primarily included patients with OSA and featured comprehensive annotations.
- **Beijing Tongren Hospital (BTH affiliated to Capital Medical University)** consists of 6,327 overnight PSG recordings, mainly from the Otorhinolaryngology Department, focusing on patients with suspected OSA.
- **Inner Mongolia Mental Health Center (IMMHC)** comprises 1,689 PSG recordings collected at its sleep center from patients diagnosed with psychiatric disorders and OSA.
- **The Second Affiliated Hospital of Soochow University (SAHSU)** comprises 72 BCG recordings from its Neurology Department. Due to missing AHI labels, 41 of these recordings were ultimately used for sleep apnea validation.
- **West China Hospital (WCH affiliated to Sichuan University)** includes 591 BCG recordings from its Sleep Medicine Center.

To ensure rigorous evaluation and mitigate data leakage, we strictly maintained patient-level separation throughout the study. Based on this principle, we adopted distinct data splitting strategies tailored to the pre-training and fine-tuning stages. For PSG Pre-training: We used 10% of full-night recordings as a development set for sleep staging. For the AHI estimation task, we selected a balanced development set containing 4,000 full-night recordings. This balanced set was used solely for internal evaluation of the PSG pre-trained model (Stage 1) to ensure it performed well across the full spectrum of AHI severity (healthy, mild, moderate, and severe). For BCG Fine-tuning and Validation: As shown in Figure 1(b), 708 full-night BCG recordings from the SSPH cohort were used for fine-tuning. Crucially, for clinical validation, we used the remaining 153 SSPH subjects (Internal Validation) and the complete WCH (591 subjects) and SAHSU (72 subjects) cohorts (External Validation). These validation cohorts retained the natural, imbalanced AHI distributions characteristic of real-world clinical populations

### 4.2 Device description

BCG recordings used in this study were acquired using the 5S Sleep Tracking Mat (Five Seasons Medical, Beijing, China), an under-pillow sensing platform designed for unobtrusive data collection in both home and clinical settings. The mat incorporates a piezoelectric BCG sensor that detects micro-vibrations arising from cardiac ejection, respiratory excursions, body movements, and snore-related airflow turbulence. Measuring 60 × 10 cm, the mat is thin and flexible enough to be fully concealed beneath a standard pillow without affecting user comfort.

As illustrated in figure 7, an external acquisition and control unit digitizes the BCG signals, performs basic preprocessing, and manages wireless communication. To enable reliable deployment across heterogeneous environments, the device supports multiple data-transfer modalities—including Bluetooth, Wi-Fi, and cellular connectivity, facilitating flexible and robust transmission of nightly recordings. The system also includes an environmental sensor module capable of measuring ambient light, temperature, and humidity; however, these environmental measurements were not utilized for model development or evaluation in the present study and are reported only to document the full sensing capabilities of the hardware platform.

**Fig. 7.**
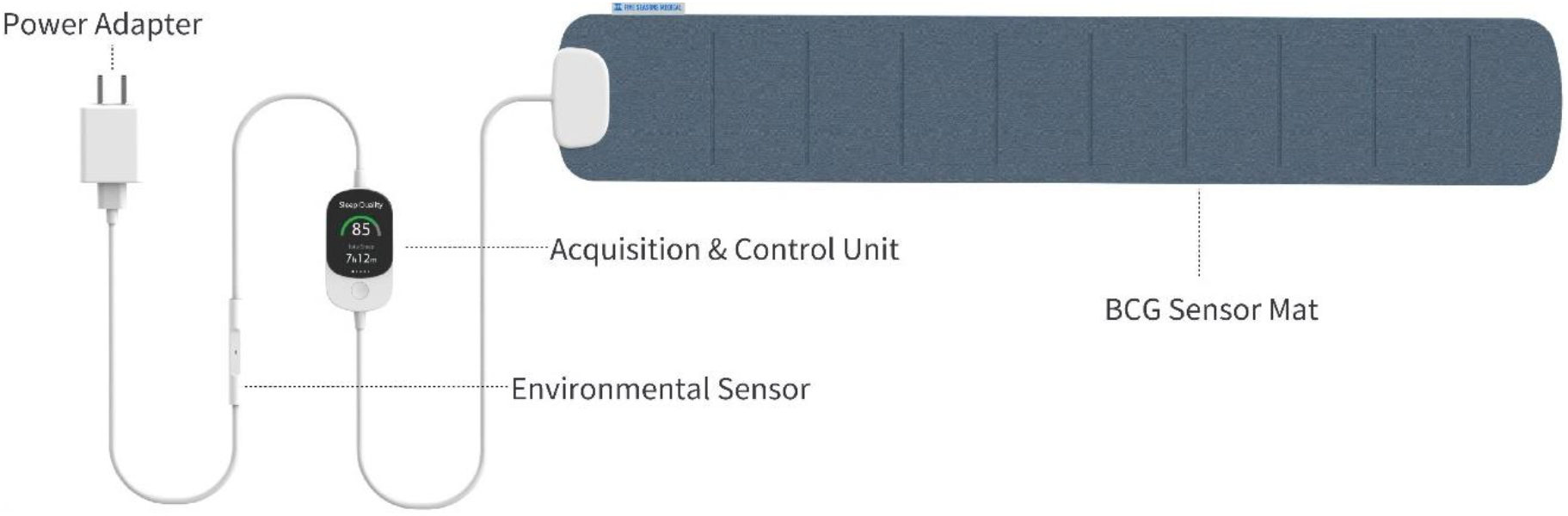
Hardware components of the 5S Sleep Tracking Mat system. The device consists of four primary modules: (1) A flexible **BCG Sensor Mat** (60 × 10 cm) placed under the pillow to capture physiological micro-vibrations; (2) An **Acquisition & Control Unit** that handles signal digitization, basic preprocessing, and data transmission; (3) An **Environmental Sensor** integrated into the power cable to monitor ambient light, temperature, and humidity; and (4) A standard **Power Adapter** for continuous operation.

### 4.3 Signal extraction

#### Signal Quality Assessment and Selection

To ensure the reliability of BCG recordings used in model fine-tuning, we implement a signal quality assessment process based on the energy distribution within respiratory and heartbeat frequency bands. Specifically, for each 15-second time segment, we compute the proportion of total signal energy contained within the respiration and heartbeat bands, denoted as the confidence score *C*:

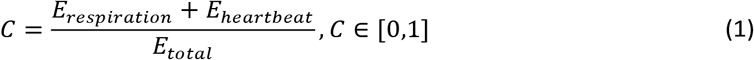

where *E*_respiration_ and *E*_heartbeat_ are the energy components in the respiration and heartbeat frequency ranges, respectively, and *E*_total_ represents the total signal energy in the segment. The 15-second window was selected to ensure the inclusion of multiple respiratory and cardiac cycles (typically 0.1–0.4 Hz and 0.6–3 Hz, respectively), allowing for a stable estimation of physiological signal presence. For each recording, an average confidence score 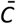 is computed over all time segments. A recording is considered valid if 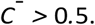 This threshold was selected based on empirical analysis: we found that recordings exceeding this level consistently retained clear physiological structure suitable for feature extraction, whereas those below it were often compromised by excessive noise or motion artifacts. Then, we process the original signal to extract the respiratory and heartbeat signals:

#### Respiratory signal extraction

The respiratory signal is derived from both the BCG and ABD signals in the PSG. First, a third-order Bessel band-pass filter [54] (0.1–0.35 Hz) is applied to the original BCG and ABD signals. The filtered signals are resampled to 4 Hz. To align the respiratory signals derived from BCG and ABD, temporal integration was applied to the BCG-derived signal. This was necessary because the BCG signal represents breathing flux, which exhibits a phase discrepancy with the volume changes measured by ABD. Finally, all signals are standardized to z-scores to eliminate magnitude differences across sensors.

#### Heartbeat signal extraction

Heartbeat intervals are represented as inter-beat intervals (IBIs). For BCG signals, J peaks define JJ intervals (JJIs), while R peaks in ECG signals represent RR intervals (RRIs). The BCG-derived heartbeats are extracted using the approach proposed by [55]. For ECG, R peaks are detected via NeuroKit2 [56]. Outliers are replaced using HRVAnalysis [57], which removes physiologically implausible intervals via linear interpolation. Removed intervals at sequence extremities are truncated. The IBIs are linearly interpolated at 4 Hz to form a continuous signal. The resulting JJIs and RRIs, expressed in milliseconds, are then converted to instantaneous heart rate (IHR) in beats per minute (BPM) via 1000 × 60*/*JJI, and the IHR are used as model inputs.

### 4.4 Model development

We propose a two-stage transfer learning model to address sleep staging and AHI3% estimation, focusing on two different data sources: PSG and BCG.

#### Stage 1 (Pre-training on PSG)

In the first stage, we train our model using two channels from PSG recordings: Heartbeat (ECG-based IHR) and Respiration (ABD belt). Each channel is fed into a dedicated 1D ResNet-like [27] feature extractor, which consists of: 1) A convolutional layer (kernel size = 7, padding = 3), followed by batch normalization and ReLU activation. 2) A max pooling layer (kernel size = 3, padding = 1). and 3) Four sequential residual blocks (each containing two ResBlock1D units). The extracted feature representations from the two channels are summed element-wise to form a combined feature. We prepend a learnable CLS token to this tensor and feed the resulting sequence into a RoFormer [28] with an embedding size of 512. A final fully connected layer produces the logits for sleep staging or AHI3% estimation. This stage learns robust representations of the heartbeat and respiratory signals from the PSG domain.

#### Stage 2 (Fine-tuning on BCG)

In this stage, we fine-tune the pre-trained model on BCG. The network processes two types of input streams with distinct sampling strategies. The Transferred channels (BCG-derived IHR and Respiration) are resampled to 4 Hz to match the input requirements of the PSG-pretrained layers, allowing the model to leverage the pretrained weights directly via *Channel-Wise Adaptation*. In contrast, the BCG-specific channels retain different sampling rates to preserve high-fidelity morphological details: BCG Respiration (0.1–1 Hz band, sampled at 2 Hz), BCG Heartbeat (0.5–25 Hz band, sampled at 50 Hz), and BCG Snore (30–200 Hz band, sampled at 400 Hz). To accommodate these varying sampling rates, we employ adaptive stride settings within the feature extractors. The token length for each channel is calculated based on the task duration (1 s for AHI3% estimation, 30 s for sleep staging) and the channel’s sampling rate. The strides in the five-block CNN extractors are configured to down-sample these inputs into feature vectors of consistent dimensions. Detailed configurations of token lengths and stride arrays for each channel are provided in Supplementary Table 6. Subsequently, a learnable gating mechanism called *Dynamic Feature Fusion* adaptively weighs the three BCG-specific feature streams, which are then fused with the two transferred channels via element-wise addition to form a comprehensive representation. This fused representation is passed through the RoFormer encoder and the final fully-connected layer. It is worth noting that while the CNN extractors capture local features, the RoFormer backbone processes the full sequence of features. This architecture enables the model to capture long-range temporal dependencies such as low-frequency heart rate variability, beyond the local receptive field of the CNNs.

#### Hyperparameters

We use the Adam optimizer with an initial learning rate of 1 × 10^*−*4^ for both tasks, reduced to 1 × 10^*−*5^ during fine-tuning. Weighted Cross Entropy is adopted for sleep staging. For AHI estimation, we utilize Focal Loss [58], where apnea and hypopnea events are grouped into one class, and normal breathing events as another. Table 5 summarizes the key training hyperparameters.

**Table 5:** Key training hyperparameters for both tasks.

| Setting | Sleep Staging | AHI3% Estimation |
| --- | --- | --- |
| Optimizer | Adam | Adam |
| Loss function | Weighted Cross Entropy Loss | Focal Loss |
| Batch size | 32 | 32 |
| Token length | 30 s | 1 s |
| Input length | Whole night | 30 min |

#### Input Segmentation and Inference Logic

For sleep staging, the entire night recording is split into consecutive 30-s segments for training and evaluation. For AHI3% estimation, we utilize a 30-minute input window during the training phase to allow the model to learn contextual event patterns efficiently. However, during the inference phase, the model slides this 30-minute window across the entire night recording to form a whole-night event sequence. The final output is a patient-level AHI3%, computed strictly according to clinical standards: the total count of predicted events divided by the TST (in hours).

### 4.5 Evaluations

For sleep staging, we use Cohen’s kappa (*κ*), Accuracy (Acc), and F1, as overall performance indicators, while Precision and Recall are reported for each sleep stage and presented in a confusion matrix.

- *κ* accounts for chance agreement between predictions and ground truth:

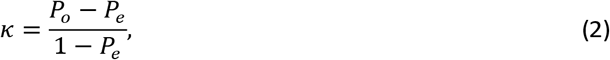

where *P*_*o*_ is the observed agreement and *P*_*e*_ is the expected agreement under random chance.
- **Acc** measures the overall proportion of correctly classified sleep stages:

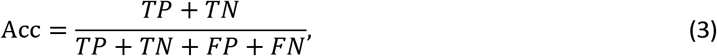

where *TP, TN, FP*, and *FN* denote the counts of true positives, true negatives, false positives, and false negatives, respectively.
- **Precision** for each sleep stage quantifies the fraction of epochs predicted as that stage that are truly of that stage:

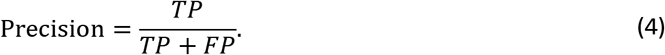
- **Recall** for each sleep stage indicates the fraction of actual epochs of that stage that are correctly identified:

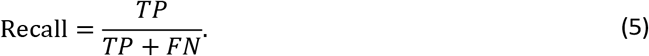
- **F1**: For each sleep stage *i*, the F1 is the harmonic mean of its precision and recall

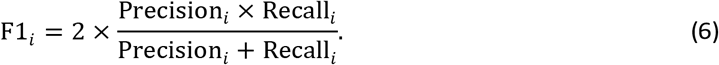

In this paper, we used the Macro-F1 as default, which computes the average of the F1-scores for each stage. This approach treats all classes equally, regardless of their prevalence in the dataset:

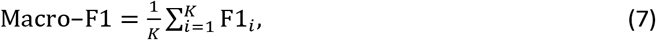

where *K* is the number of sleep stages.

We report Sensitivity (Sens) and Specificity (Spec) across three AHI severity thresholds: mild–5, moderate–15, and severe–30 sleep apnea, based on standard clinical cutoffs. For global AHI estimation, we use the Pearson correlation coefficient (*r*) as the primary evaluation metric, complemented by Scatter plots and Bland–Altman plots for graphical assessment. At the 4-class classification level, we evaluate performance using macro F1 score and confusion matrices to capture class-wise precision and recall balance.

- **Sens** measures the proportion of correctly detected positive cases:

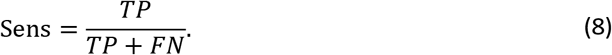
- **Spec** measures the proportion of correctly detected negative cases:

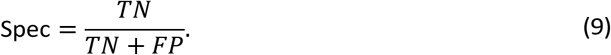
- **r** quantifies the linear correlation between the predicted AHI values and the ground truth AHI values:

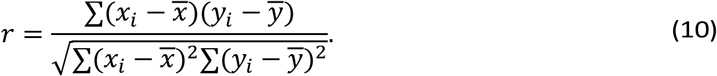

To quantify the accuracy and consistency of our model in estimating macro-architecture parameters, we evaluate the following five metrics: Total Sleep Time (TST), Sleep Onset Latency (SOL), Wake After Sleep Onset (WASO), Sleep Efficiency (SE), and REM Latency (REML). We assess each of the five sleep-quality measures with Pearson *r* and ICC to provide a complete picture of linear agreement and measurement reliability.

- **Intraclass Correlation Coefficient (ICC):**

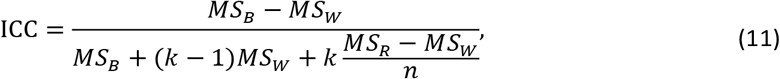

where *MS*_*B*_, *MS*_*W*_, and *MS*_*R*_ are the between-subjects, within-subjects, and residual mean squares, respectively, *k* is the number of measurements per subject (here *k* = 2, the true and predicted values), and *n* is the number of subjects.

## Data Availability

The datasets from NSRR are obtained via https://sleepdata.org, and the HSP datasets are available at https://bdsp.io/content/hsp/2.0. The Chinese cohorts are available from the authors upon reasonable request.

## Declaration statements

### 2. Code availability

The codes are open source and available at https://github.com/FiveSeasonsMedical/BCGNet.

## 3. Acknowledgements

The authors gratefully acknowledge the National Sleep Research Resource (NSRR; sleepdata.org), funded by the National Heart, Lung, and Blood Institute (NHLBI), for providing access to de-identified clinical data that supported the development and evaluation of this work. The authors also acknowledge the Brain Data Science Platform (BDSP) for access to the Human Sleep Project (HSP) dataset, a growing collection of de-identified clinical polysomnography recordings from Massachusetts General Hospital. In addition, the authors thank the clinical and research teams at Shanghai Sixth People’s Hospital (SSPH), Beijing Tongren Hospital (BTH), Inner Mongolia Mental Health Center (IMMHC), the Second Affiliated Hospital of Soochow University (SAHSU), and West China Hospital (WCH) for providing de-identified clinical datasets from Chinese cohorts that were essential for the development and validation of this work.

## 4. Author contributions

Conception and design of the study: S.C., X.C., C.L., H.Y., X.T., and Y.L.; Project supervision and administration: R.Z., D.L., C.L., Y.L., S.P., A.C., and M.B.W.; Provision of clinical resources: W.H., Y.S., F.L., X.X., D.L., and S.Y.; Algorithm design and development: S.C., X.C., C.S., Z.J., Y.W., and C.Z.; Manuscript drafting: S.C., X.C., S.H., and Y.L.; All authors revised the manuscript and approved the final version.

## References

[1] Lim, D. C. et al. The need to promote sleep health in public health agendas across the globe. Lancet Public Health 8, e820–e826 (2023).

[2] Irwin, M. R. Why sleep is important for health: a psychoneuroimmunology perspective. Annu. Rev. Psychol. 66, 143–172 (2015).

[3] Mukherjee, S. et al. An official American Thoracic Society statement: the importance of healthy sleep. recommendations and future priorities. Am. J. Respir. Crit. Care Med. 191, 1450–1458 (2015).

[4] Leng, Y., Musiek, E. S., Hu, K., Cappuccio, F. P. & Yaffe, K. Association between circadian rhythms and neurodegenerative diseases. Lancet Neurol. 18, 307–318 (2019).

[5] Rundo, J. V. & Downey III, R. Polysomnography. Handb. Clin. Neurol. 160, 381–392 (2019).

[6] Bloch, K. E. Polysomnography: a systematic review. Technol. Health Care 5, 285–305 (1997).

[7] Boulos, M. I. et al. Normal polysomnography parameters in healthy adults: a systematic review and meta-analysis. Lancet Respir. Med. 7, 533–543 (2019).

[8] Portier, F. et al. Evaluation of home versus laboratory polysomnography in the diagnosis of sleep apnea syndrome. Am. J. Respir. Crit. Care Med. 162, 814–818 (2000).

[9] Miettinen, T. et al. Home polysomnography reveals a first-night effect in patients with low sleep bruxism activity. J. Clin. Sleep Med. 14, 1377–1386 (2018).

[10] Tschopp, S., Wimmer, W., Caversaccio, M., Borner, U. & Tschopp, K. Night-to-night variability in obstructive sleep apnea using peripheral arterial tonometry: a case for multiple night testing. J. Clin. Sleep Med. 17, 1751–1758 (2021).

[11] Caples, S. M., Anderson, W. M., Calero, K., Howell, M. & Hashmi, S. D. Use of polysomnography and home sleep apnea tests for the longitudinal management of obstructive sleep apnea in adults: an American Academy of Sleep Medicine clinical guidance statement. J. Clin. Sleep Med. 17, 1287–1293 (2021).

[12] Birrer, V., Elgendi, M., Lambercy, O. & Menon, C. Evaluating reliability in wearable devices for sleep staging. NPJ Digit. Med. 7, 74 (2024).

[13] Sadek, I., Biswas, J. & Abdulrazak, B. Ballistocardiogram signal processing: a review. Health Inf. Sci. Syst. 7, 10 (2019).

[14] Kim, C.-S. et al. Ballistocardiogram: Mechanism and potential for unobtrusive cardiovascular health monitoring. Sci. Rep. 6, 31297 (2016).

[15] Yu, B., Chen, Y., Cai, D. & Zhang, H. A proof-of-concept study on non-contact BCG-based cardiac monitoring for in-patients with sleep apnea syndrome using piezoelectric ceramics. IEEE Trans. Instrum. Meas. (2025).

[16] Paalasmaa, J., Waris, M., Toivonen, H., Leppäkorpi, L. & Partinen, M. Unobtrusive online monitoring of sleep at home, 3784–3788 (IEEE, 2012).

[17] Sadek, I., Heng, T. T. S., Seet, E. & Abdulrazak, B. A new approach for detecting sleep apnea using a contactless bed sensor: Comparison study. J. Med. Internet Res. 22, e18297 (2020).

[18] Yi, R., Enayati, M., Keller, J. M., Popescu, M. & Skubic, M. Non-invasive in-home sleep stage classification using a ballistocardiography bed sensor, 1–4 (IEEE, 2019).

[19] Migliorini, M. et al. Automatic sleep staging based on ballistocardiographic signals recorded through bed sensors, 3273–3276 (IEEE, 2010).

[20] Wiard, R. M. et al. Automatic detection of motion artifacts in the ballistocardiogram measured on a modified bathroom scale. Med. Biol. Eng. Comput. 49, 213–220 (2011).

[21] Vanderperren, K. et al. Removal of BCG artifacts from EEG recordings inside the MR scanner: a comparison of methodological and validation-related aspects. Neuroimage 50, 920–934 (2010).

[22] Mitsukura, Y., Sumali, B., Nagura, M., Fukunaga, K. & Yasui, M. Sleep stage estimation from bed leg ballistocardiogram sensors. Sensors 20, 5688 (2020).

[23] Wang, Z. et al. Assessing the severity of sleep apnea syndrome based on ballistocardiogram. PLoS One 12, e0175351 (2017).

[24] Berry, R. B. et al. AASM scoring manual updates for 2017 (version 2.4) (2017).

[25] Edouard, P. et al. Validation of the Withings sleep analyzer, an under-the-mattress device for the detection of moderate-severe sleep apnea syndrome. J. Clin. Sleep Med. 17, 1217–1227 (2021).

[26] Ding, F. et al. Polysomnographic validation of an under-mattress monitoring device in estimating sleep architecture and obstructive sleep apnea in adults. Sleep medicine 96, 20–27 (2022).

[27] He, K., Zhang, X., Ren, S. & Sun, J. Deep residual learning for image recognition, 770–778 (2016).

[28] Su, J. et al. Roformer: Enhanced transformer with rotary position embedding. Neurocomputing 568, 127063 (2024).

[29] Korkalainen, H. et al. Deep learning enables sleep staging from photoplethysmogram for patients with suspected sleep apnea. Sleep 43, zsaa098 (2020).

[30] Sun, H. et al. Sleep staging from electrocardiography and respiration with deep learning. Sleep 43, zsz306 (2020).

[31] Sridhar, N. et al. Deep learning for automated sleep staging using instantaneous heart rate. NPJ Digit. Med. 3, 106 (2020).

[32] Radha, M. et al. A deep transfer learning approach for wearable sleep stage classification with photoplethysmography. NPJ Digit. Med. 4, 135 (2021).

[33] Li, S. et al. SleepNetZero: Zero-burden zero-shot reliable sleep staging with neural networks based on ballistocardiograms. Proc. ACM Interact. Mob. Wearable Ubiquitous Technol. 8, 1–25 (2024).

[34] Liu, M.-H. et al. EfficientNet-based machine learning architecture for sleep apnea identification in clinical single-lead ECG signal data sets. BioMed. Eng. OnLine 23, 57 (2024).

[35] Zhang, Y. et al. Deep learning for obstructive sleep apnea detection and severity assessment: A multimodal signals fusion multiscale transformer model. Nat. Sci. Sleep 1–15 (2025).

[36] He, H. et al. What radio waves tell us about sleep! Sleep 48, zsae187 (2025).

[37] Rao, S., Ali, A. E. & Cesar, P. Deepsleep: a ballistocardiographic deep learning approach for classifying sleep stages, 187–190 (2019).

[38] Raphelson, J. R. et al. Evaluation of a novel device to assess obstructive sleep apnea and body position. J. Clin. Sleep Med. 19, 1643–1649 (2023).

[39] Goetz, L., Seedat, N., Vandersluis, R. & van der Schaar, M. Generalization—a key challenge for responsible ai in patient-facing clinical applications. NPJ Digit. Med. 7, 126 (2024).

[40] Cohen, J. P. et al. Problems in the deployment of machine-learned models in health care. Cmaj 193, E1391–E1394 (2021).

[41] Sadek, I., Heng, T. T. S., Seet, E. & Abdulrazak, B. A new approach for detecting sleep apnea using a contactless bed sensor: Comparison study. J. Med. Internet Res. 22, e18297 (2020).

[42] Chen, X. et al. Non-contact detection of sleep apnea using multi-timescale cardiopulmonary features, 439–446 (2024).

[43] Sanchez Gomez, J., Pramono, R. X. A., Imtiaz, S. A., Rodriguez-Villegas, E. & Valido Morales, A. Validation of a wearable medical device for automatic diagnosis of OSA against standard PSG. J. Clin. Med. 13, 571 (2024).

[44] Davies, C. et al. A single-arm, open-label, multicenter, and comparative study of the ANNE sleep system vs polysomnography to diagnose obstructive sleep apnea. J. Clin. Sleep Med. 18, 2703–2712 (2022).

[45] Jaworski, D. & Park, E. J. Nonlinear heart rate variability analysis for sleep stage classification using integration of ballistocardiogram and apple watch. Nat. Sci. Sleep 1075–1090 (2024).

[46] Kwon, H. B. et al. Attention-based LSTM for non-contact sleep stage classification using IR-UWB radar. IEEE J. Biomed. Health Inf. 25, 3844–3853 (2021).

[47] Pini, N. et al. An automated heart rate-based algorithm for sleep stage classification: Validation using conventional polysomnography and an innovative wearable electrocardiogram device. Front. Neurosci. 16, 974192 (2022).

[48] Milton, S. et al. Five-year changes in 24-hour sleep-wake activity and dementia risk in oldest old women. Neurology 104, e213403 (2025).

[49] Li, P. et al. Daytime napping and Alzheimer’s dementia: a potential bidirectional relationship. Alzheimers Dement. 19, 158–168 (2023).

[50] Rosenberg, R. S. & Van Hout, S. The American Academy of Sleep Medicine inter-scorer reliability program: sleep stage scoring. J. Clin. Sleep Med. 9, 81–87 (2013).

[51] Chinoy, E. D. et al. Performance of seven consumer sleep-tracking devices compared with polysomnography. Sleep 44, zsaa291 (2021).

[52] Lee, Y. J., Lee, J. Y., Cho, J. H. & Choi, J. H. Interrater reliability of sleep stage scoring: a meta-analysis. J. Clin. Sleep Med. 18, 193–202 (2022).

[53] Rosenberg, R. S. & Van Hout, S. The American Academy of Sleep Medicine inter-scorer reliability program: sleep stage scoring. J. Clin. Sleep Med. 9, 81–87 (2013).

[54] Thomson, W. Delay networks having maximally flat frequency characteristics. Proc. IEE-Part III: Radio Commun. Eng. 96, 487–490 (1949).

[55] Brüser, C., Winter, S. & Leonhardt, S. Robust inter-beat interval estimation in cardiac vibration signals. Physiol. Meas. 34, 123 (2013).

[56] Makowski, D. et al. Neurokit2: A python toolbox for neurophysiological signal processing. Behav. Res. Methods 1–8 (2021).

[57] Massaro, S. & Pecchia, L. Heart rate variability (HRV) analysis: A methodology for organizational neuroscience. Organ. Res. Methods 22, 354–393 (2019).

[58] Lin, T.-Y., Goyal, P., Girshick, R., He, K. & Dollar, P. Focal loss for dense object detection (2017).

