## Supplement for "BCGNet: An AI Model Trained on 600K Hours of Sleep Data for a Novel Under-Pillow Contactless Monitoring Device"

The authors provide this Supplement to give readers additional information about their work.

### Supplementary results

#### Detailed Characteristics of Chinese Cohorts

**Supplementary Table 1**: Sleep characteristics of the Chinese cohorts.

| Dataset | AHI3% | Wake | N1 | N2 | N3 | REM |
|---|---|---|---|---|---|---|
| **PSG** | | | | | | |
| BTH | 24.65 ± 27.16 | 0.16 ± 0.14 | 0.10 ± 0.09 | 0.50 ± 0.16 | 0.09 ± 0.11 | 0.16 ± 0.07 |
| SSPH | 32.98 ± 28.12 | 0.18 ± 0.15 | 0.15 ± 0.12 | 0.44 ± 0.14 | 0.13 ± 0.10 | 0.10 ± 0.06 |
| IMMHC | 12.69 ± 18.28 | 0.23 ± 0.16 | 0.15 ± 0.09 | 0.49 ± 0.16 | 0.03 ± 0.05 | 0.11 ± 0.07 |
| **BCG** | | | | | | |
| SSPH[‡] | 21.45 ± 19.02 | 0.26 ± 0.15 | 0.13 ± 0.08 | 0.34 ± 0.10 | 0.10 ± 0.09 | 0.13 ± 0.05 |
| SSPH[§] | 33.80 ± 27.50 | 0.22 ± 0.13 | 0.15 ± 0.11 | 0.32 ± 0.11 | 0.11 ± 0.09 | 0.14 ± 0.06 |
| WCH | 23.85 ± 22.65 | 0.27 ± 0.12 | 0.21 ± 0.11 | 0.37 ± 0.12 | 0.01 ± 0.03 | 0.14 ± 0.06 |
| SAHSU | 25.05 ± 21.30 | 0.28 ± 0.16 | 0.12 ± 0.09 | 0.36 ± 0.11 | 0.11 ± 0.07 | 0.13 ± 0.06 |

Note: All values are presented as mean ± standard deviation. Sleep stage values represent proportions of total sleep time. The SSPH set indicated by [‡] was used for fine-tuning, while the SSPH set indicated by [§] is the internal validation set. These PSG recordings from the Chinese cohorts were used exclusively for pretraining. WCH and SAHSU served as external validation cohorts.

Supplementary Table 1 summarizes the key characteristics of the Chinese cohorts used in this study. This includes the large-scale PSG cohorts (BTH, SSPH, IMMHC) for pre-training and the independent BCG cohorts used for fine-tuning (SSPH) and validation (SSPH, WCH, SAHSU). The table details the mean Apnea-Hypopnea Index (AHI3%) and the proportional distribution of sleep stages, highlighting the diversity in sleep apnea severity and sleep architecture across the datasets.

#### Comprehensive performance of BCGNet in sleep staging

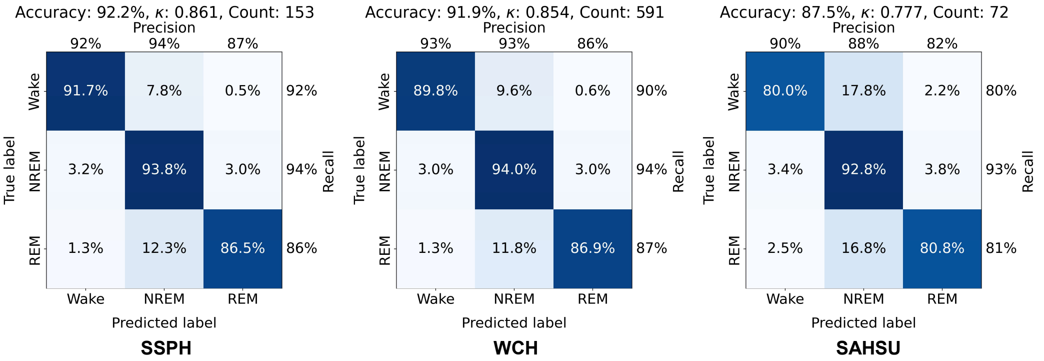


**Supplementary Figure 1**: Performance of BCGNet for 3-class sleep staging across multi-center cohorts. Performance is reported for the internal validation cohort (SSPH), and the external validation cohorts (WCH and SAHSU).

To further characterize the sleep staging capabilities of BCGNet, we evaluated its performance on both 3-class (Wake, NREM, REM) and 5-class (Wake, N1, N2, N3, REM) classifications across three independent, multi-center cohorts (SSPH, WCH, and SAHSU). The results, detailed in Supplementary Figure 1 and 2, demonstrate the BCGNet's robust and highly accurate performance. For the 3-class task, BCGNet achieved outstanding overall accuracies of 92.2%, 91.9%, and 87.5%, with corresponding Cohen's $\kappa$ of 0.861, 0.854, and 0.777, respectively. Moreover, the results on the external cohorts indicate a very high level of agreement (e.g., Cohen's $\kappa$ of 0.854 in the WCH) with gold-standard PSG scoring and underscore the BCGNet's reliability in distinguishing fundamental sleep stages.

In the more granular and challenging 5-class staging task, BCGNet continued to exhibit strong performance, attaining accuracies of 78.3%, 74.4%, and 74.0% ($\kappa = 0.714$, 0.654, and 0.655) across the

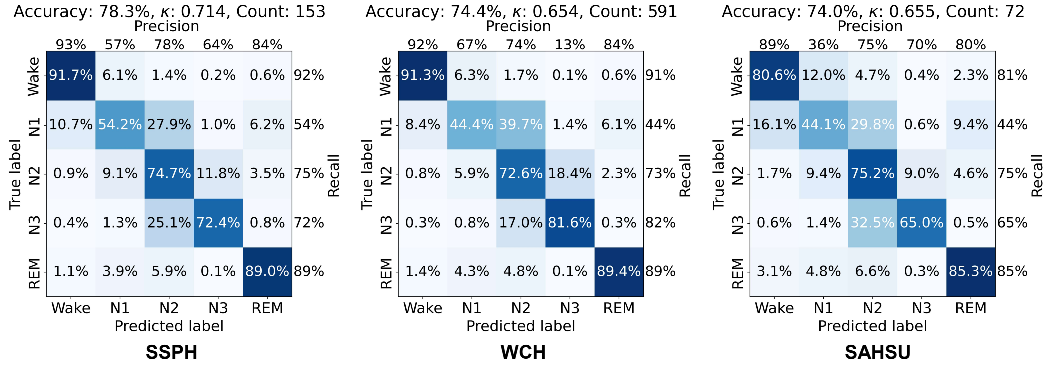


**Supplementary Figure 2**: Performance of BCGNet for 5-class sleep staging across multi-center cohorts. Performance is reported for the internal validation cohort (SSPH), and the external validation cohorts (WCH and SAHSU).

SSPH, WCH, and SAHSU cohorts. The confusion matrix of the external cohorts (Accuracy: 74.3%, $\kappa$: 0.655) reveals that the BCGNet excels at identifying clinically significant stages, such as deep sleep (N3) and REM sleep. While minor confusion exists between adjacent, lighter sleep stages (e.g., N1 and N2), a well-known challenge even for human experts, the overall high performance confirms BCGNet's accuracy for detailed sleep architecture analysis across diverse populations.

#### Robust Classification of Sleep Apnea Severity

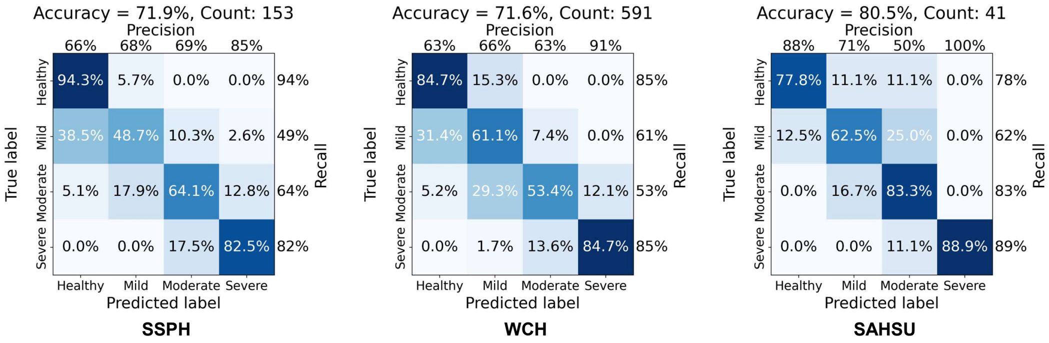


**Supplementary Figure 3**: Performance of BCGNet for AHI3% classification (Healthy, Mild, Moderate, Severe) across multi-center cohorts. Performance is reported for the internal validation cohort (SSPH) and the external validation cohorts (WCH and SAHSU).

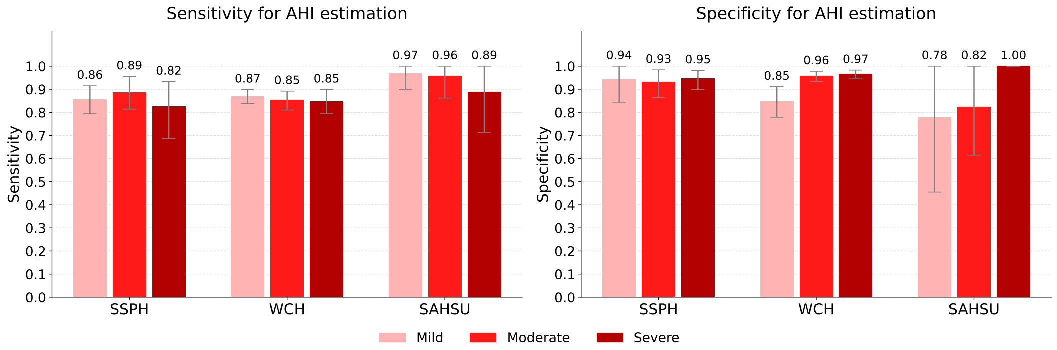


**Supplementary Figure 4**: Performance of BCGNet at standard AHI cutoffs. AHI cutoffs of 5, 15, and 30 were used to classify mild, moderate, and severe cases, respectively. Performance is reported for the internal set (SSPH), WCH, SAHSU, and the external cohort (WCH and SAHSU).

We rigorously assessed BCGNet's ability to classify obstructive sleep apnea (OSA) severity based on the AHI3%. This evaluation was conducted using both a four-category classification (Healthy, Mild, Moderate, Severe) and standard clinical cutoffs. As shown in Supplementary Figure 3, BCGNet achieved high overall accuracy in the four-class task across all three validation cohorts (71.9% in SSPH, 71.6% in WCH, and 75.0% in SAHSU). Notably, the BCGNet demonstrated superior recall for severe cases (82%, 85%, 89%). This highlights the BCGNet's utility in reliably detecting patients with the most severe form of the disease.

The clinical viability of BCGNet is further substantiated by its performance at the standard AHI cutoffs of 5, 15, and 30 events/hr (Supplementary Figure 4). The BCGNet exhibited consistently high sensitivity and specificity across all thresholds and cohorts. For instance, specificity for severe OSA (AHI $> 30$) was exceptional, reaching 0.95 (SSPH), 0.97 (WCH), and a perfect 1.00 (SAHSU), indicating a near-zero false positive rate for severe diagnoses. Concurrently, sensitivity for detecting moderate-to-severe disease remained high, affirming that BCGNet is a powerful tool for screening and severity assessment in clinical practice.

**Supplementary Table 2**: Sensitivity and specificity of BCGNet on 3-class sleep staging.

| Metrics | SSPH | WCH | SAHSU | External Cohorts |
|---|---|---|---|---|
| Wake-Sens | 0.917 (0.904,0.930) | 0.898 (0.888,0.907) | 0.800 (0.769,0.833) | 0.889 (0.879,0.897) |
| NREM-Sens | 0.938 (0.930,0.946) | 0.940 (0.935,0.943) | 0.928 (0.911,0.944) | 0.938 (0.935,0.942) |
| REM-Sens | 0.865 (0.838,0.888) | 0.869 (0.857,0.879) | 0.808 (0.754,0.855) | 0.862 (0.851,0.873) |
| Wake-Spec | 0.972 (0.965,0.977) | 0.973 (0.970,0.975) | 0.968 (0.956,0.979) | 0.972 (0.969,0.975) |
| NREM-Spec | 0.907 (0.895,0.918) | 0.897 (0.890,0.904) | 0.826 (0.798,0.851) | 0.890 (0.882,0.896) |
| REM-Spec | 0.978 (0.974,0.981) | 0.978 (0.976,0.979) | 0.967 (0.959,0.974) | 0.977 (0.975,0.978) |

Sens = sensitivity; Spec = specificity. Performance is reported for the internal cohort (SSPH), WCH, SAHSU, and the external cohort (WCH and SAHSU).

**Supplementary Table 3**: Sensitivity and specificity of BCGNet on 4-class sleep staging.

| Metrics | SSPH | WCH | SAHSU | External Cohorts |
|---|---|---|---|---|
| Wake-Sens | 0.912 (0.897, 0.925) | 0.903 (0.893, 0.912) | 0.785 (0.746, 0.824) | 0.891 (0.881,0.901) |
| Light-Sens | 0.838 (0.824, 0.853) | 0.797 (0.790, 0.805) | 0.847 (0.826, 0.869) | 0.801 (0.795,0.808) |
| Deep-Sens | 0.662 (0.617, 0.709) | 0.867 (0.828, 0.903) | 0.611 (0.554, 0.665) | 0.732 (0.693,0.772) |
| REM-Sens | 0.874 (0.847, 0.897) | 0.870 (0.858, 0.881) | 0.822 (0.775, 0.864) | 0.865 (0.853,0.877) |
| Wake-Spec | 0.972 (0.966, 0.977) | 0.971 (0.968, 0.974) | 0.966 (0.952, 0.979) | 0.971 (0.968,0.974) |
| Light-Spec | 0.860 (0.844, 0.874) | 0.902 (0.894, 0.908) | 0.775 (0.746, 0.803) | 0.886 (0.878,0.893) |
| Deep-Spec | 0.954 (0.947, 0.962) | 0.919 (0.916, 0.923) | 0.970 (0.962, 0.976) | 0.924 (0.920,0.927) |
| REM-Spec | 0.975 (0.971, 0.978) | 0.976 (0.974, 0.978) | 0.966 (0.957, 0.974) | 0.975 (0.973,0.977) |

Sens = sensitivity; Spec = specificity. Performance is reported for the internal cohort (SSPH), WCH, SAHSU, and the external cohort (WCH and SAHSU).

**Supplementary Table 4**: Sensitivity and specificity of BCGNet on 5-class sleep staging.

| Metrics | SSPH | WCH | SAHSU | External Cohorts |
|---|---|---|---|---|
| Wake-Sens | 0.917 (0.902, 0.930) | 0.913 (0.905, 0.921) | 0.806 (0.771, 0.838) | 0.903 (0.894,0.910) |
| N1-Sens | 0.542 (0.504, 0.580) | 0.444 (0.428, 0.460) | 0.441 (0.392, 0.492) | 0.444 (0.429,0.459) |
| N2-Sens | 0.747 (0.727, 0.764) | 0.726 (0.715, 0.737) | 0.752 (0.725, 0.778) | 0.729 (0.719,0.738) |
| N3-Sens | 0.724 (0.673, 0.772) | 0.816 (0.763, 0.863) | 0.650 (0.576, 0.722) | 0.729 (0.684,0.775) |
| REM-Sens | 0.890 (0.864, 0.911) | 0.894 (0.883, 0.904) | 0.853 (0.807, 0.895) | 0.890 (0.879,0.900) |
| Wake-Spec | 0.974 (0.969, 0.978) | 0.970 (0.966, 0.973) | 0.963 (0.948, 0.977) | 0.969 (0.966,0.972) |
| N1-Spec | 0.937 (0.928, 0.944) | 0.943 (0.939, 0.946) | 0.917 (0.903, 0.930) | 0.940 (0.936,0.943) |
| N2-Spec | 0.883 (0.870, 0.895) | 0.849 (0.842, 0.857) | 0.856 (0.836, 0.877) | 0.850 (0.842,0.858) |
| N3-Spec | 0.951 (0.943, 0.959) | 0.928 (0.923, 0.932) | 0.960 (0.952, 0.968) | 0.930 (0.926,0.935) |
| REM-Spec | 0.973 (0.969, 0.977) | 0.974 (0.972, 0.976) | 0.962 (0.954, 0.969) | 0.973 (0.971,0.975) |

Sens = sensitivity; Spec = specificity. Performance is reported for the internal cohort (SSPH), WCH, SAHSU, and the external cohort (WCH and SAHSU).

Supplementary Table 2, 3, and 4 show the performances of BCGNet in 3-, 4-, and 5- sleep staging using sensitivity and specificity. The results in 3-class staging show strong performance of BCGNet across three sleep stages (Wake, NREM, and REM). Its high sensitivity and specificity values, particularly for NREM and REM, indicate reliable sleep staging, although there is a slight dip in performance for the SAHSU cohort. For the 4-class task, BCGNet achieved consistently high wake and REM sensitivity and specificity across all three cohorts, with slightly reduced sensitivity in the deep stage, particularly in SAHSU. In terms of the 5-class task, performance remained strong for wake, N2, N3, and REM, but sensitivity for N1 was markedly lower across all datasets, consistent with the known difficulty of distinguishing stage N1. Overall, the model demonstrated robust generalizability across cohorts with stable specificity.

#### High agreement in sleep continuity and architecture parameter estimation

Beyond sleep staging and AHI3% estimation, we validated BCGNet's capacity to derive key parameters of sleep continuity and architecture. We conducted Bland-Altman analyses to compare BCGNet-predicted values against PSG-derived ground truth for five standard metrics: Total Sleep Time (TST), Sleep Onset Latency (SOL), Wake After Sleep Onset (WASO), Sleep Efficiency (SE), and REM Latency (REML). The results, presented in Supplementary Figure 5, reveal a remarkable level of agreement across all three independent cohorts.

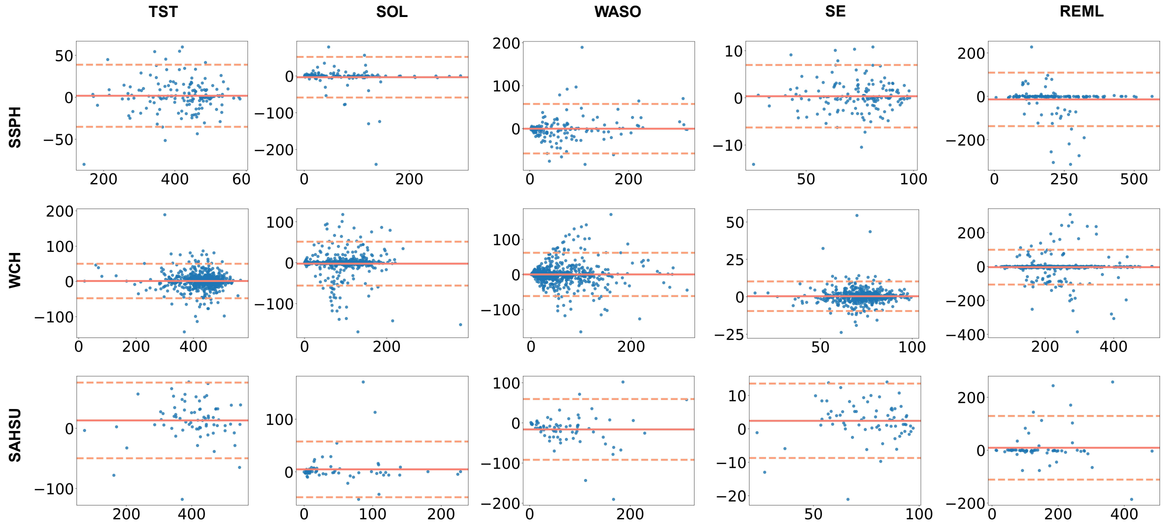


**Supplementary Figure 5**: Bland-Altman plots for overnight sleep continuity and architecture parameter estimation. This figure illustrates the agreement between BCGNet predicted values and ground-truth expert annotations for five sleep continuity and architecture parameters (TST, SOL, WASO, SE, REML) in the cohorts: SSPH, WCH, and SAHSU. Each panel shows the difference between predicted and true values against their mean. The solid red line represents the mean bias, and the dashed red lines indicate the 95% LoA.

For all five parameters, the mean bias (solid red line) remained near zero for most cases, suggesting that the BCGNet generally does not exhibit a strong systematic over- or under-estimation. Furthermore, the 95% limits of agreement (dashed red lines) were narrow, demonstrating strong concordance at the individual subject level. These results underscore BCGNet's potential as a reliable and comprehensive tool for sleep analysis, capable of capturing nuanced aspects of sleep quality and structure from BCG signals alone.

#### Ablation study on external cohorts

Supplementary Table 5 summarizes the ablation results on external cohorts. In both the WCH and SAHSU datasets, BCGNet achieved superior performance compared with the BCG-based baseline, with consistent improvements in both sleep staging and AHI3% estimation. Notably, while PSG-based models without fine-tuning also performed strongly, BCGNet either matched or surpassed them, particularly in the SAHSU cohort where it yielded the highest staging agreement ($\kappa$ = 0.697) and F1 score (0.781). These results highlight the effectiveness of BCGNet in adapting to heterogeneous external cohorts and its robustness in both sleep staging and AHI3% estimation tasks.

**Supplementary Table 5**: Ablation study on sleep staging and AHI3% estimation in the external cohorts.

| Method | 4-class sleep staging | | AHI3% estimation | |
|---|---|---|---|---|
| | **Cohen's $\kappa$** | **F1** | **Pearson's $r$** | **F1** |
| *WCH Cohort* | | | | |
| BCG-based | 0.646 (0.636, 0.655) | 0.644 (0.634, 0.653) | 0.947 (0.933, 0.959) | 0.680 (0.639, 0.717) |
| PSG-based (*w/o* fine-tuning) | 0.733 (0.725, 0.741) | 0.707 (0.697, 0.715) | 0.952 (0.940, 0.961) | 0.678 (0.639, 0.717) |
| **BCGNet (proposed)** | **0.736 (0.728, 0.743)** | **0.710 (0.700, 0.718)** | **0.959 (0.949, 0.968)** | **0.703 (0.668, 0.741)** |
| *SAHSU Cohort* | | | | |
| BCG-based | 0.546 (0.513, 0.581) | 0.665 (0.641, 0.693) | 0.933 (0.887, 0.972) | 0.679 (0.517, 0.813) |
| PSG-based (*w/o* fine-tuning) | 0.687 (0.655, 0.715) | 0.771 (0.748, 0.790) | 0.935 (0.894, 0.972) | **0.766 (0.595, 0.886)** |
| **BCGNet (proposed)** | **0.697 (0.666, 0.725)** | **0.781 (0.759, 0.800)** | **0.952 (0.920, 0.981)** | 0.764 (0.595, 0.892) |

"*w/o*" stands for "without". The results are based on the external WCH and sAHSU cohorts. The bolded values indicate the highest performance for each task.

#### Adaptive Temporal Down-sampling Strategy

To effectively fuse physiological signals recorded at vastly different sampling rates, ranging from 2 Hz for respiration to 400 Hz for snore audio, we designed a modality-specific input processing pipeline. The goal was to preserve the high-frequency morphological details of the raw signals while ensuring that the extracted feature vectors from all encoders possess consistent temporal dimensions for the subsequent *Dynamic Feature Fusion* module.

As detailed in Supplementary Table 6, the token length ($L_{in}$) for each channel is determined by the product of the task-specific window duration ($T$) and the channel's native sampling rate ($f_s$): $L_{in} = T \times f_s$. For the AHI3% estimation task ($T$ = 1s) and the Sleep Staging task ($T$ = 30s), this results in input vectors of varying lengths. To align these inputs, the 1D ResNet feature extractor for each channel employs a customized configuration of convolutional strides across its five blocks. The product of the strides approximates the input length, effectively down-sampling the temporal dimension to a unified scale.

**Supplementary Table 6**: Detailed configuration of each signal modality.

| Signal Modality | Sampling Rate (Hz) | AHI3% Estimation (Token duration = 1 s) | | Sleep Staging (Token duration = 30 s) | |
|---|---|---|---|---|---|
| | | **Token Length** | **Strides (5 Blocks)** | **Token Length** | **Strides (5 Blocks)** |
| *Transferred Channels* | | | | | |
| PSG/BCG Resp/Heart | 4 | 4 | [1, 1, 1, 2, 2] | 120 | [2, 2, 2, 3, 5] |
| *BCG-Specific Channels* | | | | | |
| BCG Respiration | 2 | 2 | [1, 1, 1, 1, 2] | 60 | [1, 2, 2, 3, 5] |
| BCG Heartbeat | 50 | 50 | [1, 1, 2, 5, 5] | 1500 | [2, 3, 5, 5, 5] |
| BCG Snore | 400 | 400 | [2, 4, 4, 5, 5] | 12000 | [4, 4, 4, 5, 15] |

The "Strides" column lists the stride values applied sequentially in the five blocks of the ResNet encoder. The distinct stride combinations ensure that despite the varying input lengths (Token Length), the output feature maps are down-sampled to a compatible temporal resolution for feature fusion.
